# Do established comorbidity scores predict in-hospital mortality equally well in women and men? A nationwide validation in 166.7 million German inpatient cases, 2010–2024

**DOI:** 10.64898/2026.09.14.26362826

**Authors:** Josua A. Decker, Hamed Mirbagheri, Simon Hellbrueck, Lisa Maria Pfadenhauer, Ute Seeland, Thomas Kroencke, Christian Scheurig-Muenkler

## Abstract

**Objectives:** To determine whether three established comorbidity scores (Charlson Comorbidity Index, Elixhauser Comorbidity Sum, van Walraven Score), applied with a single formula to both sexes, are equally well calibrated and equally discriminating in women and men, and whether any difference reaches a pre-specified threshold of clinical relevance.

**Design:** Population-based retrospective cohort study; external validation of three pre-existing prediction models (TRIPOD Type 4) with sex-stratified evaluation. The analysis plan was fixed in the time-stamped Research Data Centre submission syntax before any aggregated data existed.

**Setting:** Germany; complete national enumeration of acute somatic Diagnosis-Related Group (DRG)-billed adult inpatient cases, reporting years 2010 to 2024.

**Participants:** 166 693 491 adult inpatient cases meeting a pre-specified nine-step eligibility filter.

**Main outcome measures:** In-hospital mortality. Primary axis: calibration (calibration-in-the-large, CITL; calibration slope; observed mortality per score quintile). Secondary axis: discrimination (area under the receiver operating characteristic curve, AUC). Pre-specified thresholds: between-sex slope difference of at least 0.05, absolute AUC difference of at least 0.01.

**Results:** Calibration-in-the-large was effectively perfect in both sexes (|CITL| ≤ 0.0004). The scores diverged in how prediction kept pace with rising risk, and in opposite directions for women and men: for the Charlson index the calibration slope was 0.968 in men and 1.034 in women (difference 0.066), with smaller differences for the Elixhauser-Sum (0.039) and the van Walraven Score (0.029). At the same score quintile, observed mortality was higher in men at most quintiles, with a maximum risk ratio of 1.21 (95 % CI 1.20 to 1.22); two van Walraven quintiles ran the other way (0.87 and 0.97). Differences in discrimination were small (random-effects AUC difference: Charlson −0.0076, 95 % CI −0.0095 to −0.0057; Elixhauser-Sum +0.0074; van Walraven −0.0026).

**Conclusions:** Applied with a single formula to both sexes, three established comorbidity scores are calibrated differently in women and men, whereas their discrimination differs only marginally. Because these scores are reused at scale in research and in risk-adjusted mortality comparisons between hospitals, a systematic difference in calibration does not average out. This supports sex-specific recalibration. The downstream effect on hospital benchmarking is not yet quantified and should not be presumed negligible.

**Study registration:** OSF DOI 10.17605/OSF.IO/P3QAW.

**Plain Language Summary:** Comorbidity scores allow for cross-hospital mortality comparisons for how ill patients already were on admission, by combining their pre-existing conditions into a single risk number. Worldwide, three scores are predominantly used: The Charlson Comorbidity Index, the Elixhauser Comorbidity Sum, and the van Walraven Score. All three are applied to men and women using the same formula. Only the van Walraven Score was originally built to predict death in hospital. The Charlson index was built for long-term survival, and the Elixhauser Sum is a simple unweighted count of conditions. None had been checked separately for men and women in a large population.

We applied the three scores to 166.7 million adult hospital stays in Germany between 2010 and 2024, selected from the national hospital record by pre-specified inclusion criteria. Separately for men and women, we analysed two things: Whether the predicted risk matched the mortality actually observed (calibration), and how well the score separated patients who died in hospital from those who survived (discrimination). Calibration differed systematically between the sexes. At the same score, men with the same predicted risk in fact died more often than women, by up to about a fifth across risk groups, and the scores tracked rising risk at slightly different rates in men and women, in opposite directions. Differences in how well the scores told apart those who died from those who survived were very small between the sexes, smaller than a thousandth on a scale that runs from 0.5 to 1.

For an individual patient these differences are too small to change a treatment decision. They matter because these scores are used everywhere, to compare the mortality of whole hospitals and in thousands of research studies, and a score that fits one sex better than the other bends those comparisons in a consistent direction. A small bias that always points the same way, repeated across millions of cases, does not cancel out. This is why the calibration differences, not the very small discrimination differences, are the result that counts, and why their full effect still needs to be measured. The pattern held across all 15 years and across age groups.

**What this paper adds:**

- Comorbidity scores derived from hospital discharge data are applied to men and women with a single single formula for both sexes, although sex-related differences in calibration of the Charlson index has been reported for long-term outcomes. No nationwide evaluation of all three of the most widely used scores by sex existed for in-hospital mortality.
- In a sex-stratified validation on 166 693 491 German adult inpatient cases (2010– 2024), all three scores were systematically miscalibrated by sex when applied with a single formula to both sexes: At equal scores men had higher in-hospital mortality than women (risk ratios up to 1.21), while discrimination differed only marginally. The pattern held across all 15 years and across the age range, and reversed between emergency and elective admissions.
- Because these scores are reused at scale in hospital benchmarking and observational research, a systematic sex bias in their calibration does not average out but is reproduced wherever they are applied with a single formula to both sexes. This justifies investigating sex-specific recalibration, and the downstream impact should not be presumed negligible.

## Introduction

Comorbidity scores derived from administrative hospital discharge data are the standard instrument for adjusting in-hospital mortality for how ill patients already were on admission. Their use extends well beyond the individual prognosis they were built for: They enter standardised mortality ratio (SMR) computations as continuous values, underpin public hospital-quality reporting and, in several health systems, reimbursement, and increasingly feed clinical decision-support algorithms. Sex and gender have been documented as systematically under-considered axes in clinical research [1], and cardiovascular risk prediction has responded by moving to sex-specific models such as the QRISK3 algorithms [2] and the AHA PREVENT equations [3]. Risk adjustment has not followed: the instruments in routine use apply one formula to men and women alike. Throughout this paper we use sex in the biological and administrative meaning in which it is recorded in the German Diagnosis-Related Group statistics, that is, as a binary variable documented at the case level, and we distinguish it from gender as the sociocultural dimension of roles, behaviour and treatment within the health system, following SAGER 2016 [4]. Gender is not represented in these data; our results therefore speak to sex as recorded and support no inference about gender identity or gender-related care pathways. A sex-neutral score applied to a sex-stratified population does not only distort individual risk estimates, it can also advantage or disadvantage hospitals systematically according to the sex composition of their caseload, independent of the quality of care delivered. The regulatory and reporting environment makes this timely: The TRIPOD 2015 statement [5] and its 2024 update TRIPOD+AI [6] added explicit fairness items, and the EU AI Act 2024 [7], in full application from August 2026, requires demonstrated equity across demographic subgroups including sex for systems classed as high-risk clinical decision support.

Three scores dominate this literature. The Charlson Comorbidity Index was developed in 1987 in 559 medical inpatients with 1-year mortality as the outcome and validated over 10 years in 685 women treated for breast cancer [8]. It was re-weighted for hospital discharge abstracts by Quan and colleagues in 2011 [9]. The Elixhauser comorbidity measures were developed in 1998 on 1.8 million California discharges [10] and mapped to the International Classification of Diseases, 10th revision (ICD-10), by Quan in 2005 [11]. The variant examined here, the Elixhauser Comorbidity Sum, is the unweighted count of the 31 comorbidity domains. The van Walraven Score is a point-score adaptation of the same domains, developed in 2009 on 228 565 Ontario admissions with in-hospital death as its explicit outcome [12]. A systematic review identified 54 studies comparing these scores [13], and the Agency for Healthcare Research and Quality re-validated the Elixhauser index in 2017 on 18 US state databases [14]. How well such a score performs is not one question but several. Steyerberg and Vergouwe distinguish four axes [15]: (A) Calibration-in-the-large, whether the mean predicted risk matches the observed event rate; (B) the calibration slope, whether prediction keeps pace with risk across the range rather than being too extreme or too compressed; (C) concordance, the ability to rank patients who die above those who survive; and (D) clinical usefulness, assessed by decision curves. For risk adjustment the calibration axes carry more weight than concordance, because these scores enter SMR computations as continuous values rather than at a decision threshold: A score that ranks only moderately well but is correctly calibrated within each stratum still yields unbiased benchmarking, whereas a score that ranks well but is miscalibrated in one stratum yields biased benchmarking. The three scores also stand in different relationships to the outcome examined here. Only the van Walraven Score was built for in-hospital death, the Charlson index was built for longer-term mortality and re-purposed through the Quan re-weighting, and the Elixhauser Sum, being an unweighted count, does not weight its components by how strongly each predicts death. None of the three was validated separately in women and men at development, and the sex-stratified evidence since then covers the Charlson index alone and mostly long-term outcomes: Rius and colleagues found sex-related miscalibration of an adapted Charlson index in a Spanish population-based cohort [16], a difference located on the calibration rather than the discrimination axis, and Westerberg and colleagues showed in a Swedish nationwide cohort that the index discriminates long-term mortality well, with a c-index of approximately 0.81 that improves with a longer look-back [17]. Whether these three scores, applied with a single single formula for both sexes, are equally well calibrated and equally discriminating in men and women, and whether any difference is large enough to bias risk-adjusted hospital mortality comparisons, has not been tested at population scale. We evaluated all three in parallel on the complete enumeration of German adult inpatient cases meeting pre-specified eligibility criteria for the reporting years 2010 to 2024, with calibration pre-specified as the primary axis, discrimination as the secondary axis, and effect-size thresholds fixed in advance. We hypothesised sex-related differences in calibration and discrimination that would be stable across reporting years and age strata but sensitive to clinical context.

## Methods

### Study design and registration

This is a population-based retrospective cohort study on a full enumeration of cases, designed as an external validation of three pre-existing prediction models without any re-development of the models themselves (TRIPOD Type 4). The analysis plan, comprising the eligibility filter, the score definitions, the evaluation approach, the sensitivity sets, the age strata, the performance metrics and the pooling method, was fixed in the Research Data Centre submission syntax that was developed and submitted before any aggregated output existed; that time-stamped syntax is reproduced in Supplementary File S2. Reporting follows STROBE 2007 [18], RECORD 2015 [19] for studies based on routinely collected health data, TRIPOD 2015 Type 4 and TRIPOD+AI 2024, and SAGER 2016 [4]. The study was registered on the Open Science Framework (OSF) after the aggregated outputs had been returned and quality-checked, and the registration is therefore declared as retrospective on the platform [20].

### Data source and cohort construction

German Diagnosis-Related Group (DRG) statistics (§21 Hospital Reimbursement Act data) for reporting years 2010 through 2024 were accessed via the Research Data Centre (Forschungsdatenzentrum, FDZ) of the Federal Statistical Office and the Statistical Offices of the Federal States under Project 4509-2021 [21]. The DRG system covers acute somatic inpatient care. Psychiatric admissions (billed under the separate PEPP system), rehabilitation admissions, and certain federal and military hospitals operate under different billing frameworks and are not included. Per reporting year, raw cases were filtered through a fixed nine-step eligibility procedure (Figure 1). The nine steps in order are as follows. (1) Adult age 18 to 110 years. (2) Standard inpatient admission only, defined by German DRG admission-reason codes 1 and 2 corresponding to elective and emergency inpatient stays. (3) No inter-hospital transfer in, defined by absence of German DRG admission-origin codes V, A, or K (transfer from another hospital, from a referring physician, or from a hospital practitioner). (4) No inter-hospital transfer out, defined by absence of German DRG discharge codes 6, 8, 13, or 29 (discharge to another hospital, to rehabilitation, to inpatient psychiatric care, or to a partial-inpatient facility). (5) Length of stay at least 2 days. (6) Sex documented in the record. (7) No pregnancy or delivery as the principal diagnosis (ICD-10 chapter O). (8) No admissions for oncology therapy only, defined by the German Modification of ICD-10 (ICD-10-GM) code Z51 as the principal diagnosis. (9) No accounting-only or dummy hospital departments. The final analytic cohort comprises 166 693 491 adult cases. Year-to-year variability in per-step exclusion shares (per cent of raw admissions) is bounded. Across the 15 reporting years, the per-step range was at most 1.1 percentage points for the age, transfer-in, transfer-out, sex, pregnancy, oncology, and pseudo-department steps. The outpatient or day-case step varies by 1.1 percentage points and the short-stay step varies by 4.5 percentage points, with a rising trend from 12.9 % in 2010 to 17.4 % in 2024 that is consistent with the progressive substitution of inpatient stays by ambulant care. All step variabilities remain below the 5 percentage-point threshold typically considered material for cohort-construction stability.

**Figure 1.**
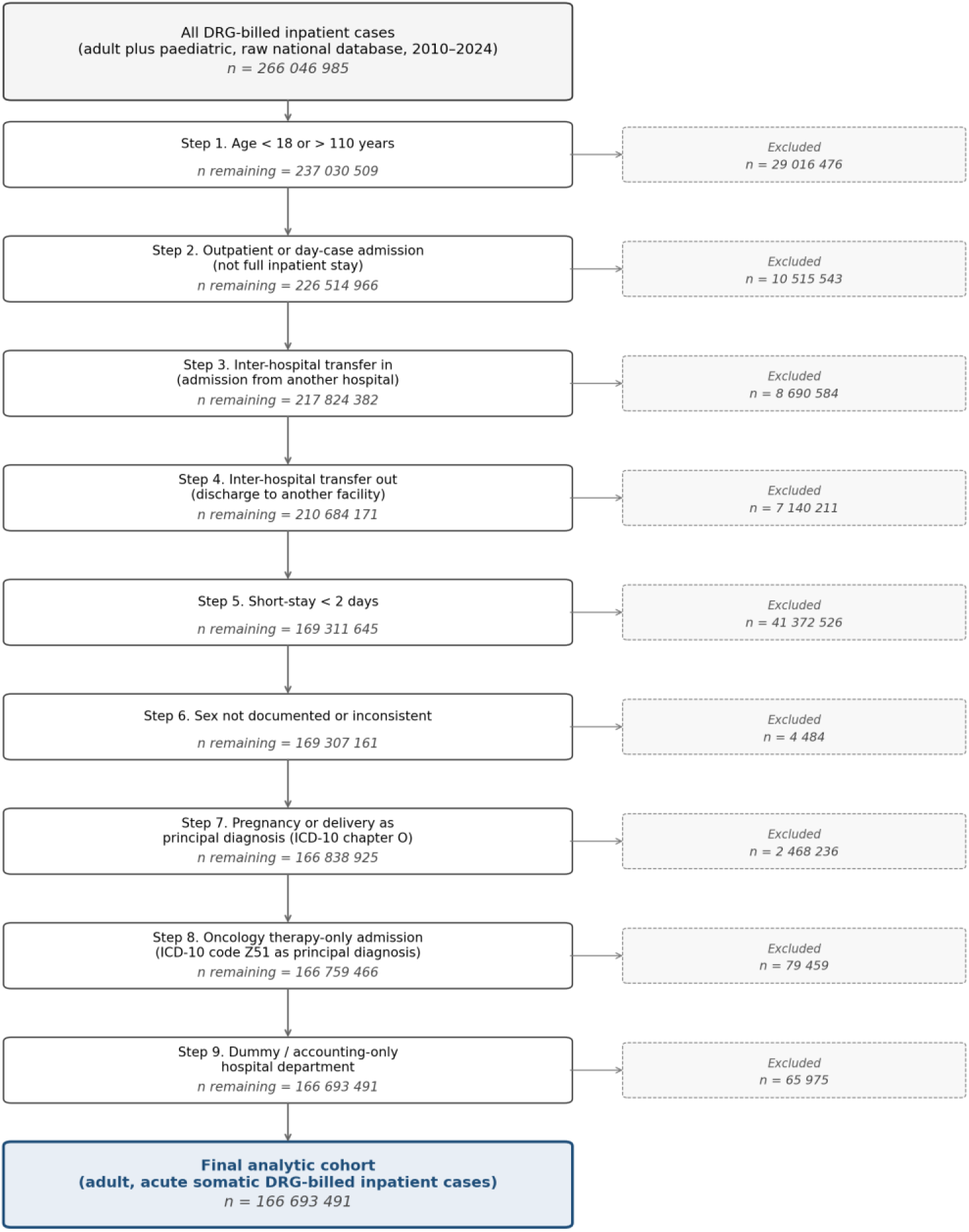
Cohort attrition flow diagram, German DRG statistics, all 15 reporting years (2010–2024). Per-step exclusion counts are direct 15-year sums from the per-year FDZ flowchart outputs. Gross-total of 266 046 985 raw admissions reduces to the final analytic cohort of 166 693 491 adult acute somatic inpatient cases through nine pre-specified exclusion steps.

### Outcome and predictors

Primary outcome: In-hospital mortality, defined as administrative discharge code death (binary, case-level). Per case, all three scores were computed from both the principal discharge diagnosis (*Hauptdiagnose*) and all secondary discharge diagnoses (*Nebendiagnosen*). Comorbidity domains were identified with the enhanced ICD-10 coding algorithm of Quan and colleagues [11], applied to the German modification ICD-10-GM. The Charlson Comorbidity Index used the weight update of Quan and colleagues [9], the Elixhauser Comorbidity Sum an unweighted count of 31 comorbidity domains [10], and the van Walraven Score the published point system [12]. The full ICD-10-GM code definitions and the component weights for all three scores are provided in Supplementary File S1. Sex is the administrative-binary variable as recorded in the DRG statistics; the term “sex” (not “gender”) is used throughout per SAGER 2016 [4].

### Statistical analysis

Per score, a pooled logistic regression without a sex term of in-hospital mortality on the score is fit on the main cohort. Predicted probabilities are evaluated in sex-stratified subgroups; this evaluation approach mirrors the operational use of the scores in national risk-adjustment frameworks (where a single a single calibration for both sexes is applied to a sex-stratified population) and detects the resulting miscalibration.

Consistent with the operational use of comorbidity scores in continuous risk-adjustment rather than at binary decision points, we pre-specified calibration as the primary axis and discrimination as the secondary axis. Calibration was quantified by calibration-in-the-large (CITL: observed minus mean-predicted mortality per sex stratum, so that a value near zero indicates predictions correct on average), the calibration slope with 95 % confidence interval (i.e. a slope below 1 indicates predicted risks that are too extreme, too high in the highest-risk patients and too low in the lowest, and a slope above 1 the reverse), Brier score (i.e. the mean squared error of the predicted probability, reported descriptively only because it is dominated by the underlying mortality rate and is therefore not comparable between sexes), the empirical decile-level observed-vs-predicted relationship, and observed in-hospital mortality per score quintile, defined as score-value quintiles of the pooled cohort with men and women compared within the same band (a non-parametric calibration check that does not depend on the linear-predictor assumption). Discrimination was quantified by the area under the receiver operating characteristic curve (AUC) with binormal standard error; sex comparison via Wald-Z for independent AUCs [22]. Inverse-variance fixed-effects pooling combines per-year effects; DerSimonian-Laird random-effects estimates [23] run in parallel and are reported as the default for confidence intervals (treating the 15 reporting years as independent realisations of the data-generating process). Between-year heterogeneity of the pooled estimates across the 15 reporting years was quantified by Higgins I² [24]. Because the complete-enumeration sample size makes every confidence interval extremely narrow and every test statistic very large (Wald-Z values reach the tens), we interpret differences by their pre-specified magnitude and by the stability of their direction across years and age strata, not by statistical significance or by the width of confidence intervals. For the same reason, between-year heterogeneity (Higgins I²) is typically very high even when the direction of an effect is stable, because minute year-to-year differences are estimated with near-certainty; we therefore read a high I² as variation in the size of an effect, not as instability in its direction.

### Pre-specified clinical-meaningfulness thresholds

Statistical significance is uninformative at our realised sample size. We pre-specified clinical-meaningfulness thresholds independent of p-values. The pre-registration (16 June 2026) specified a between-sex calibration-slope difference of at least 0.05 and an area-under-the-curve difference of at least 0.01 as substantive. The calibration-slope threshold is applied to the between-sex difference, |slope(men) − slope(women)| ≥ 0.05, consistent with the pre-registered primary hypothesis. Effects are reported and interpreted against these thresholds rather than against p-values. This approach follows the rationale of Rothman that effect-size pre-specification is the appropriate inferential discipline in large-N descriptive epidemiology, rather than nominal-p adjustment [25].

### Verification, software, reporting standards, ethics, patient and public involvement (PPI)

The meta-pooling pipeline was verified independently along three concurrent calculation paths (scipy.stats, statsmodels.stats.meta_analysis, and a bare-numpy implementation of the DerSimonian-Laird estimator), which agreed to nine decimal places. The Research Data Centre syntax was developed in Stata 17 and executed by Research Data Centre staff in Stata 19; the subsequent aggregation and meta-analysis used Python 3 with pandas, scipy.stats, statsmodels, openpyxl and matplotlib. Reporting follows STROBE 2007 [18], RECORD 2015 [19] for studies using routinely collected health data, TRIPOD 2015 [5] and TRIPOD+AI 2024 [6], and SAGER 2016 [4]; the PROBAST 2019 [26] and PROBAST+AI 2025 [27] tools were used for risk-of-bias appraisal. The completed checklists are provided as submission appendices. Ethics: The study analyses exclusively anonymised, aggregated official statistics under §27 of the German Federal Statistics Act. No patient records were inspected, no key or linkage file exists, and the authors never had access to case-level data. Research on data without personal reference is not subject to mandatory ethics committee review under German professional law. Patients were not directly involved in the design, conduct or reporting of this study, although the research question emerged from concerns raised by patient and clinician advocacy groups about algorithmic equity, and a German lay summary will be made available through the institutional website after publication. Large language models (Claude Opus 5) were used for language editing, for code review of the post-processing pipeline and for literature triage; they were not used to generate data, to select analyses or to draw conclusions, and all authors verified the content they produced.

## Results

### Cohort characteristics

The analytic cohort comprises 166 693 491 adult inpatient cases (81.2 million men, 85.5 million women; 48.7 % / 51.3 %) over 15 reporting years. Baseline characteristics by sex are reported in Table 1; key sex differences are higher prevalence of liver disease, mechanical ventilation, metastatic cancer, cardiac arrhythmia, and solid tumour in men, and higher prevalence of depression, fluid/electrolyte disorders, and sex-specific cancer as principal diagnosis in women.

**Table 1.** Cohort characteristics, German DRG statistics 2010–2024, pooled across 15 reporting years (n = 166 693 491). Weighted averages across all reporting years. Comorbidities derived from principal and secondary diagnosis codes per Quan 2005/2011 and van Walraven 2009 mappings.

| Characteristic | Men (n = 81 231 402) | Women (n = 85 462 089) |
| --- | --- | --- |
| Mean age, years | 63.9 | 66.2 |
| In-hospital mortality, % | 2.89 | 2.46 |
| Mean length of stay, days | 7.52 | 7.53 |
| Emergency admission, % | 49.1 | 48.8 |
| Mechanical ventilation, % | 2.54 | 1.67 |
| Congestive heart failure, % | 14.62 | 13.22 |
| Cardiac arrhythmia, % | 21.52 | 18.70 |
| Chronic pulmonary disease, % | 10.76 | 9.65 |
| Diabetes (uncomplicated), % | 15.51 | 13.30 |
| Chronic kidney disease, % | 14.77 | 14.11 |
| Liver disease, % | 3.94 | 2.71 |
| Metastatic cancer, % | 5.18 | 4.36 |
| Solid tumour (non-metastatic), % | 12.37 | 9.75 |
| Depression, % | 3.47 | 6.92 |
| Fluid/electrolyte disorders, % | 16.88 | 19.84 |
| Mean Charlson Comorbidity Index | 1.49 | 1.32 |
| Mean Elixhauser Comorbidity Sum | 2.36 | 2.33 |
| Mean van Walraven Score | 6.38 | 5.57 |

### Calibration by sex (primary axis)

Calibration-in-the-large was effectively perfect in both sexes for all three scores (|CITL| ≤ 0.0004 absolute mortality probability), so the sex-related component of the calibration findings is located on the slope axis, not the intercept axis (Table 2). The between-sex difference in the calibration slope is largest for the Charlson index (0.066) and smallest for the van Walraven Score (0.029). The direction of the slope deviation is opposite between Charlson and Elixhauser-Sum. Across deciles both sexes deviate from perfect calibration in a similar shape, with the between-sex difference expressed in the systematic slope of the deviation (Figure 2). This slope difference is sign-stable across all 15 reporting years (Figure 3).

**Figure 2.**
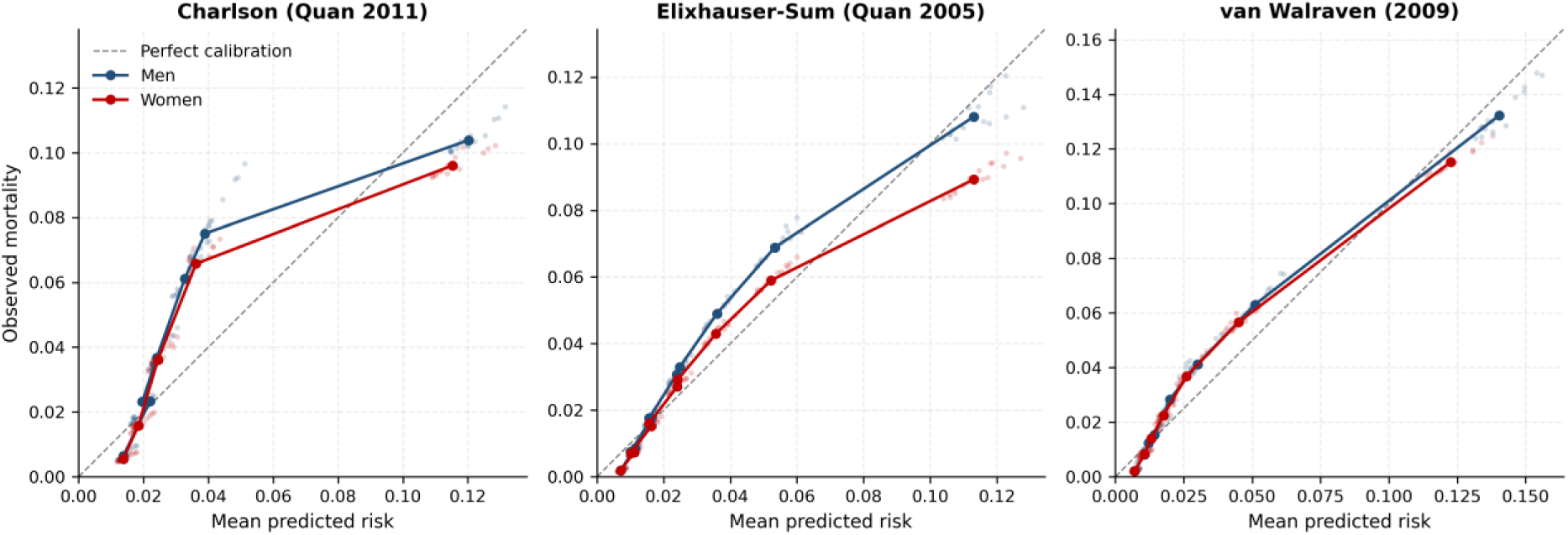
Decile calibration plots for the three scores, sex-stratified, pooled 2010–2024. Bold markers and lines show the pooled score-decile bins by sex; faint points show the individual per-year decile values (2010–2024). The dashed line marks perfect calibration. Bins are deciles of the score value, not of predicted risk; because the Charlson and van Walraven scores carry a large mass of patients at the lowest values, several low deciles collapse into a single bin, so fewer than ten points appear for these scores, whereas the Elixhauser-Sum spreads across all ten. At a given predicted risk observed mortality tends to run higher in men, and the deviation from the diagonal is greatest for the Charlson index.

**Figure 3.**
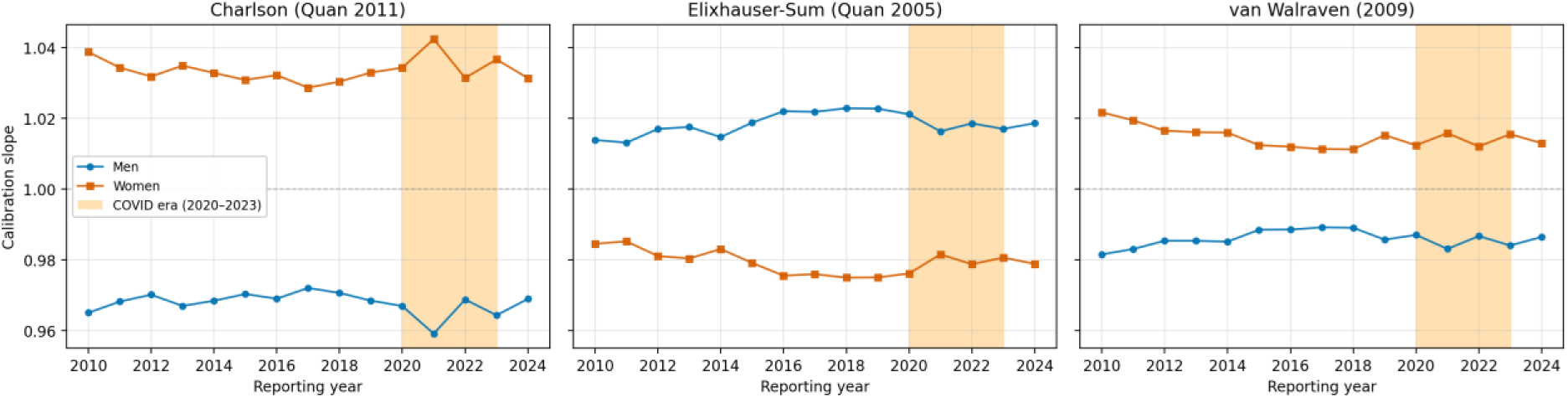
Calibration slope by reporting year, score, and sex. Reference line at 1.0 (perfect calibration). Sign-stable across 15 reporting years.

**Table 2.** Calibration of the three scores, sex-stratified, pooled 2010–2024. CITL = Calibration-in-the-large. Slope confidence intervals (CIs) from per-year DerSimonian-Laird random-effects pooling. Brier scores are descriptive only; their absolute difference between sexes is dominated by the underlying difference in outcome prevalence.

| Score | Sex | CITL (O – E) | Cal. slope (95 % CI) | Brier (descriptive) |
| --- | --- | --- | --- | --- |
| Charlson (Quan 2011) | Men | +0.0004 | 0.968 (0.965–0.971) | 0.0276 |
|  | Women | –0.0004 | 1.034 (1.031–1.037) | 0.0236 |
| Elixhauser-Sum (Quan 2005) | Men | +0.0003 | 1.018 (1.015–1.021) | 0.0274 |
|  | Women | –0.0003 | 0.979 (0.977–0.982) | 0.0237 |
| van Walraven (2009) | Men | +0.0002 | 0.986 (0.983–0.989) | 0.0267 |
|  | Women | –0.0002 | 1.015 (1.012–1.018) | 0.0230 |

### Observed mortality per score quintile by sex

Direct comparison of observed mortality at the same score quintile, stratified by sex, provides a non-parametric calibration check that does not depend on the linear-predictor assumption (Table 3, Figure 4). Men had higher observed mortality than women at most quintiles of all three scores, with risk ratios up to 1.21 (95 % CI 1.20 to 1.22; Charlson Q3); the exceptions were van Walraven Q2 and Q4, where women’s mortality was slightly higher (risk ratios 0.87, 95 % CI 0.83 to 0.90, and 0.97, 95 % CI 0.97 to 0.98).

**Figure 4.**
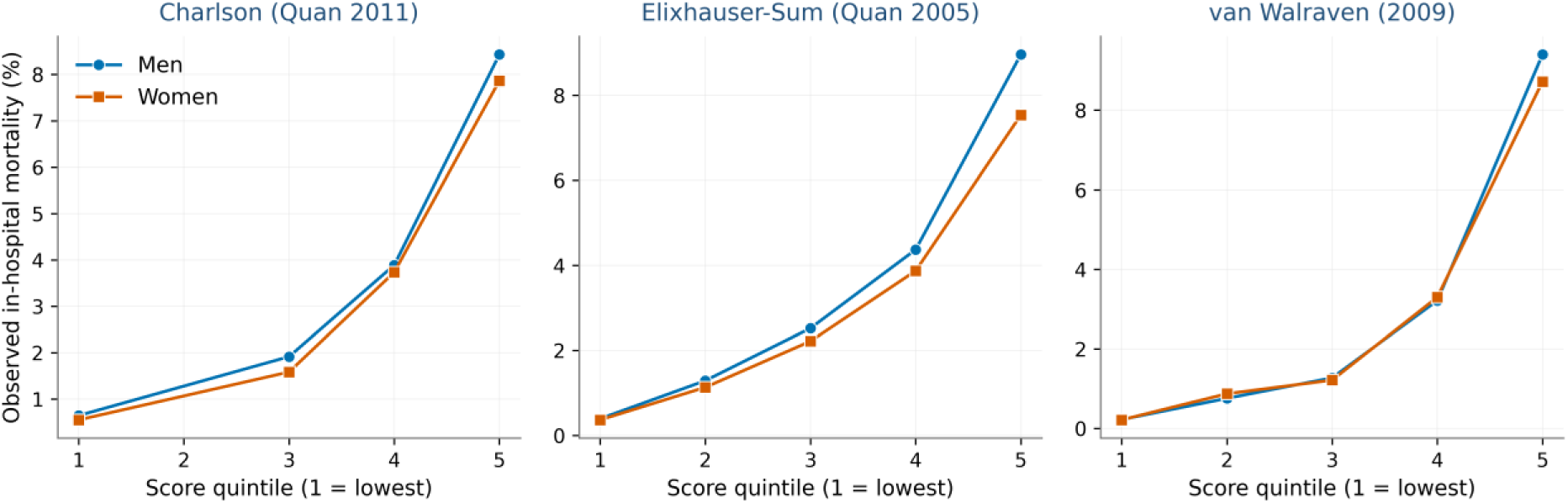
Observed in-hospital mortality per score quintile, sex-stratified, pooled across all 15 reporting years.

**Table 3.**
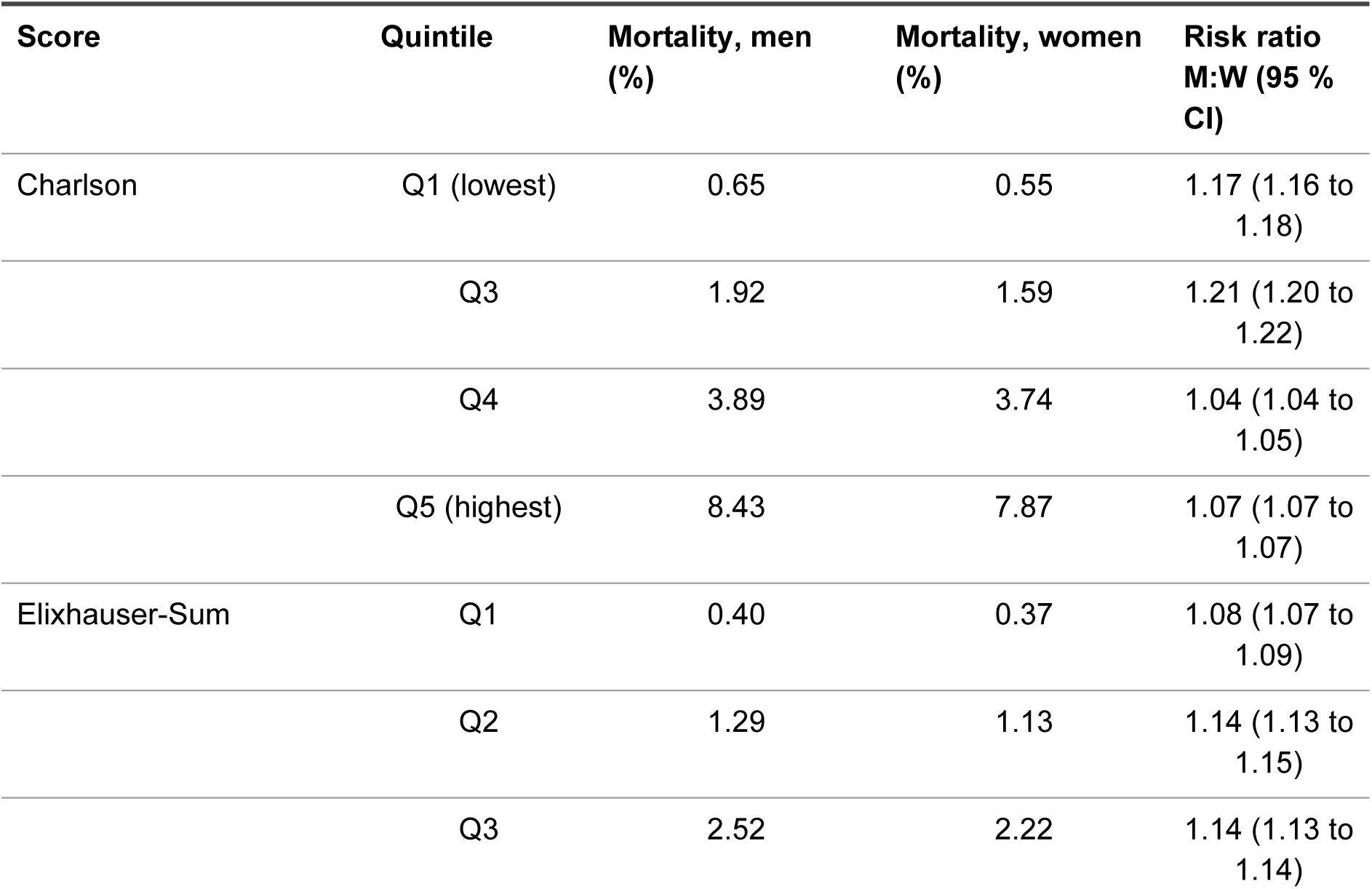
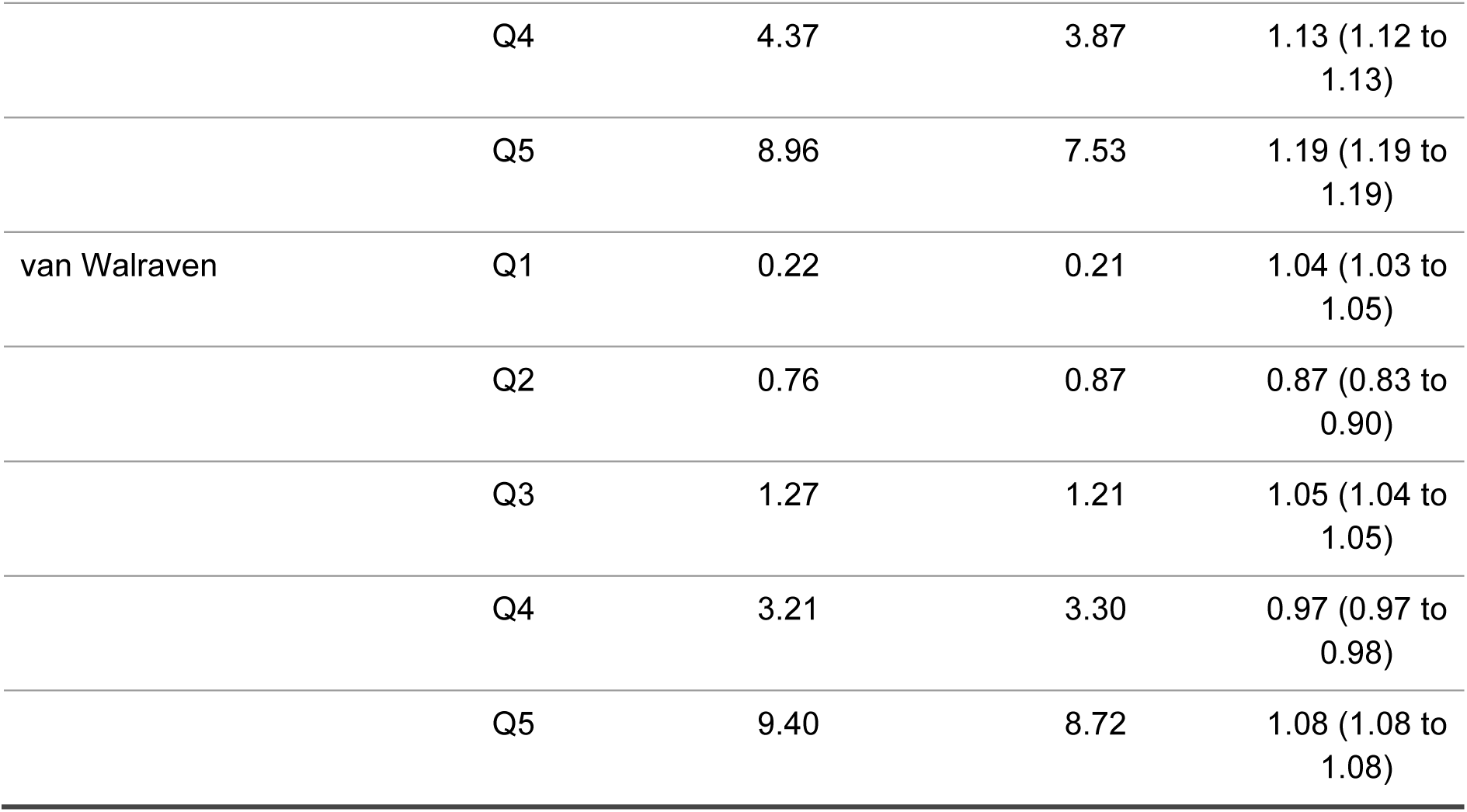
Observed in-hospital mortality per score quintile, sex-stratified, pooled 2010–2024. Risk ratio M:W = the proportion of men who died in hospital divided by the corresponding proportion in women within the same score quintile. Because the denominator is the number of cases and not person-time, these are risk ratios rather than rate ratios. Confidence intervals are Katz log-transform intervals computed from the pooled 15-year case and death counts. Quintile cut-points are determined per score by the empirical score-value distribution. For the Charlson Comorbidity Index, no separate Q2 row is reported because the dominant frequency of score = 0 in the population means the Q1 and Q2 cut-points coincide on a single score value, and the FDZ output collapses them into Q1. For the van Walraven score, the small Q2 stratum and the Q4 stratum show risk ratios below 1.0 (women mortality slightly higher than men), in contrast to the other quintiles. Because each quintile spans a range of score values, within-band differences in the score distribution between men and women can also contribute to these risk ratios; the continuous calibration slope, which does not depend on binning, is therefore the primary calibration measure. Risk ratios are computed from unrounded proportions and may therefore differ slightly from the ratio of the rounded values shown.

| Score | Quintile | Mortality, men (%) | Mortality, women (%) | Risk ratio M:W (95 % CI) |
| --- | --- | --- | --- | --- |
| Charlson | Q1 (lowest) | 0.65 | 0.55 | 1.17 (1.16 to 1.18) |
|  | Q3 | 1.92 | 1.59 | 1.21 (1.20 to 1.22) |
|  | Q4 | 3.89 | 3.74 | 1.04 (1.04 to 1.05) |
|  | Q5 (highest) | 8.43 | 7.87 | 1.07 (1.07 to 1.07) |
| Elixhauser-Sum | Q1 | 0.40 | 0.37 | 1.08 (1.07 to 1.09) |
|  | Q2 | 1.29 | 1.13 | 1.14 (1.13 to 1.15) |
|  | Q3 | 2.52 | 2.22 | 1.14 (1.13 to 1.14) |
|  | Q4 | 4.37 | 3.87 | 1.13 (1.12 to 1.13) |
|  | Q5 | 8.96 | 7.53 | 1.19 (1.19 to 1.19) |
| van Walraven | Q1 | 0.22 | 0.21 | 1.04 (1.03 to 1.05) |
|  | Q2 | 0.76 | 0.87 | 0.87 (0.83 to 0.90) |
|  | Q3 | 1.27 | 1.21 | 1.05 (1.04 to 1.05) |
|  | Q4 | 3.21 | 3.30 | 0.97 (0.97 to 0.98) |
|  | Q5 | 9.40 | 8.72 | 1.08 (1.08 to 1.08) |

### Score discrimination by sex (secondary axis)

All three between-sex discrimination differences were small (random-effects ΔAUC: Charlson −0.0076, Elixhauser-Sum +0.0074, van Walraven −0.0026) and were directionally consistent (higher discrimination in women, in men, and in women, respectively). Pooled discrimination of each score by sex is reported in Table 4; the sex-difference in discrimination and its direction in Table 5. The direction of the discrimination difference is stable across 15 reporting years for all three scores, with high between-year heterogeneity in magnitude (I² 85–96 %) but a consistent direction (Figure 5). The between-sex difference on the calibration slope is roughly 5 to 11 times the between-sex difference in discrimination (the ratio of the slope difference to the AUC difference: Charlson 8.7, Elixhauser-Sum 5.3, van Walraven 11.2; the two measures are on different scales, so this expresses relative signal size rather than a like-for-like comparison).

**Figure 5.**
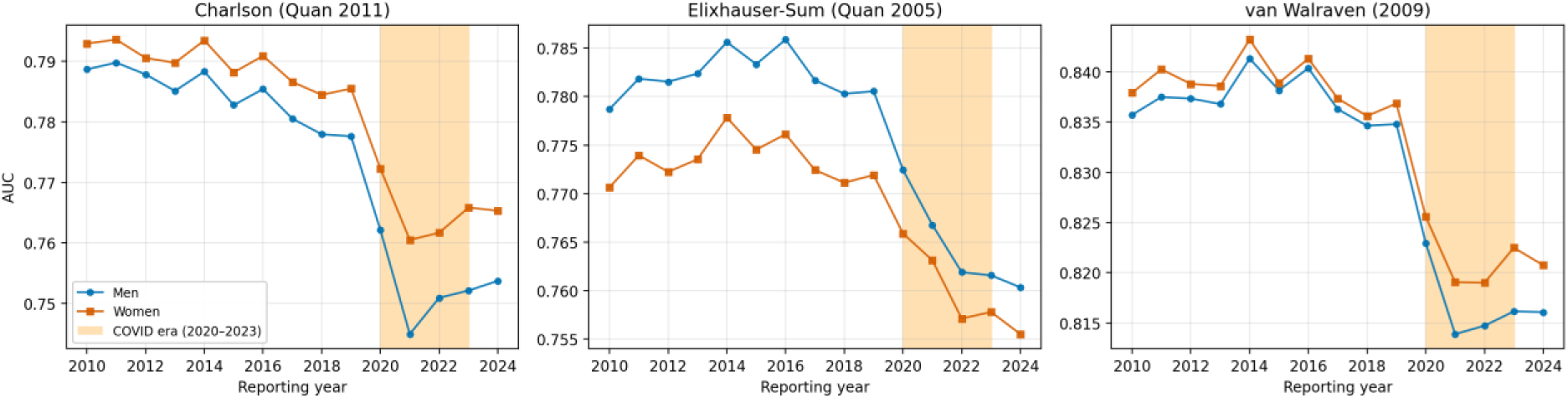
Area under the receiver operating characteristic curve by reporting year, score, and sex. COVID era (2020 to 2023) shaded.

**Table 4.** Pooled discrimination by sex, main cohort 2010–2024. Pooled AUC obtained by DerSimonian-Laird random-effects meta-analysis across 15 reporting years; the 95 % confidence intervals reflect between-year heterogeneity (I² shown).

| Score | Sex | Pooled AUC (95 % CI) | $I^2$ (%) | AUC range |
| --- | --- | --- | --- | --- |
| Charlson (Quan 2011) | Men | 0.7739 (0.7657–0.7820) | 99.9 | 0.745–0.790 |
|  | Women | 0.7814 (0.7752–0.7877) | 99.8 | 0.761–0.794 |
| Elixhauser-Sum (Quan 2005) | Men | 0.7763 (0.7717–0.7809) | 99.7 | 0.760–0.786 |
|  | Women | 0.7689 (0.7653–0.7725) | 99.4 | 0.756–0.778 |
| van Walraven (2009) | Men | 0.8305 (0.8252–0.8357) | 99.8 | 0.814–0.841 |
|  | Women | 0.8331 (0.8286–0.8375) | 99.7 | 0.819–0.843 |

**Table 5.**
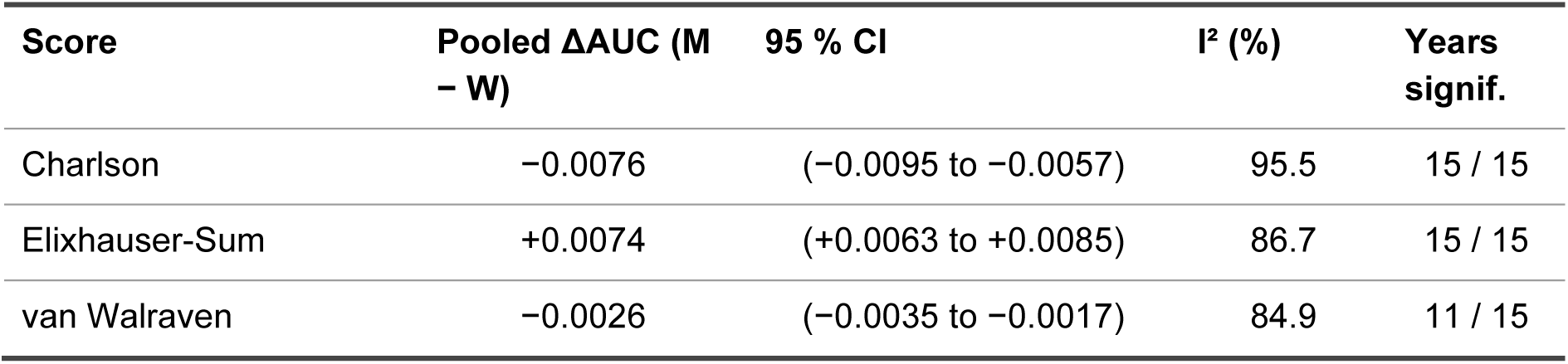
Pooled AUC difference (men − women) per score, main analysis (DerSimonian-Laird random-effects). Years significant: Number of the 15 reporting years in which the per-year difference reached statistical significance; the direction of the difference was concordant across all 15 years for every score. Higgins I² quantifies between-year heterogeneity in effect magnitude; the direction is sign-stable across all 15 years for Charlson and Elixhauser-Sum, and across 11 of 15 years for van Walraven.

### Sensitivity analyses

The seven pre-specified analysis sets for the van Walraven score (Table 6, Figure 6) show three patterns. First, restricting the cohort attenuates the sex difference in discrimination progressively: Excluding breast and prostate cancer leaves it essentially unchanged (ΔAUC −0.0021 versus −0.0025 in the main analysis), excluding all sex-specific cancers brings it to neutrality (−0.0001), and excluding all sex-specific diagnoses reverses it to favour men (+0.0017). Second, re-including pregnancy-related cases widens the difference in the original direction (−0.0066). Third, the largest contrast is between admission types, where emergency admissions favour men (+0.0077) and elective admissions favour women (−0.0114); both are large relative to the main estimate. The admission-type sets are subgroup contrasts rather than robustness checks in the strict sense, although they were pre-specified together with the other sensitivity sets.

**Figure 6.**
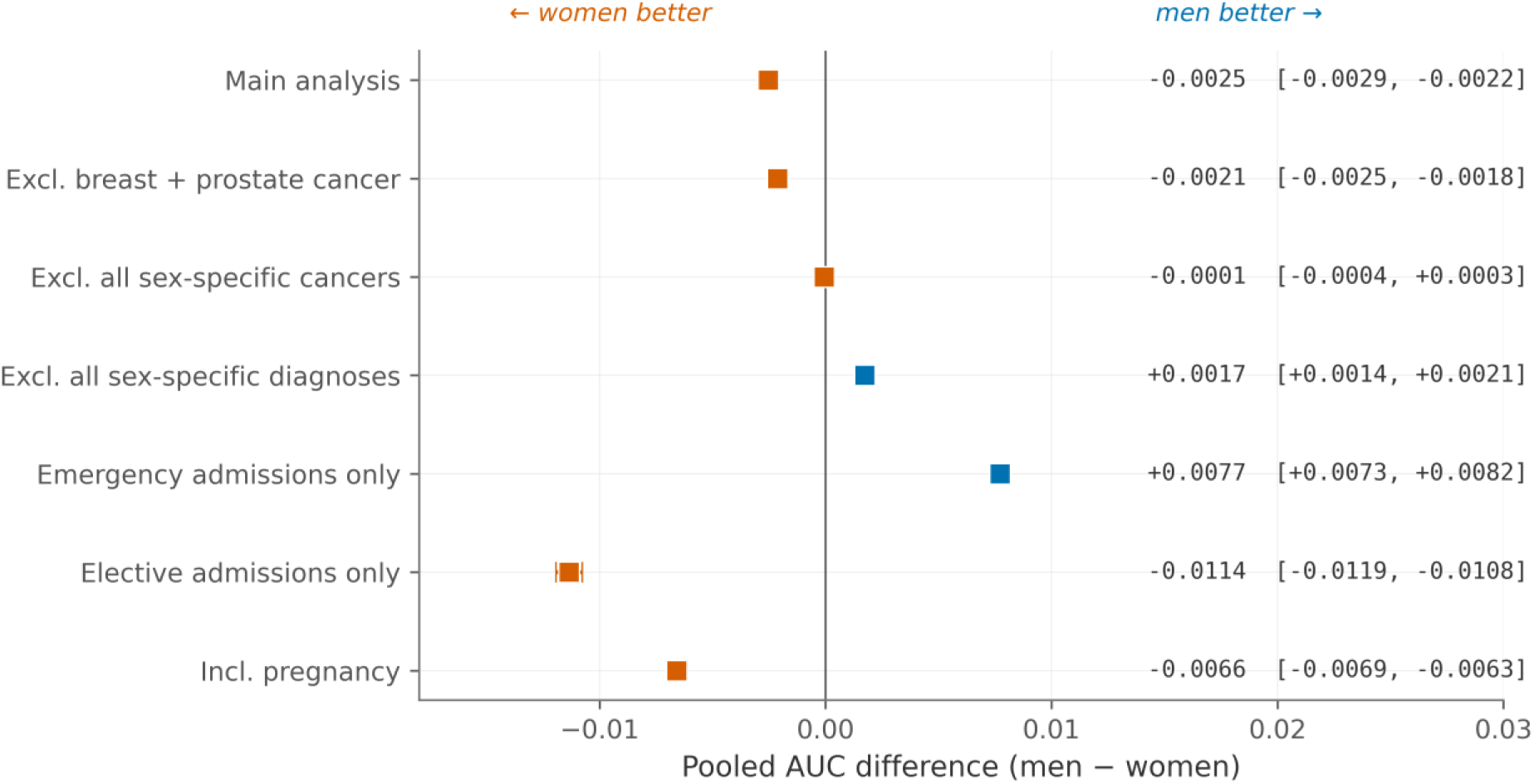
Forest plot of meta-pooled ΔAUC for the van Walraven score across seven sensitivity sets.

**Table 6.**
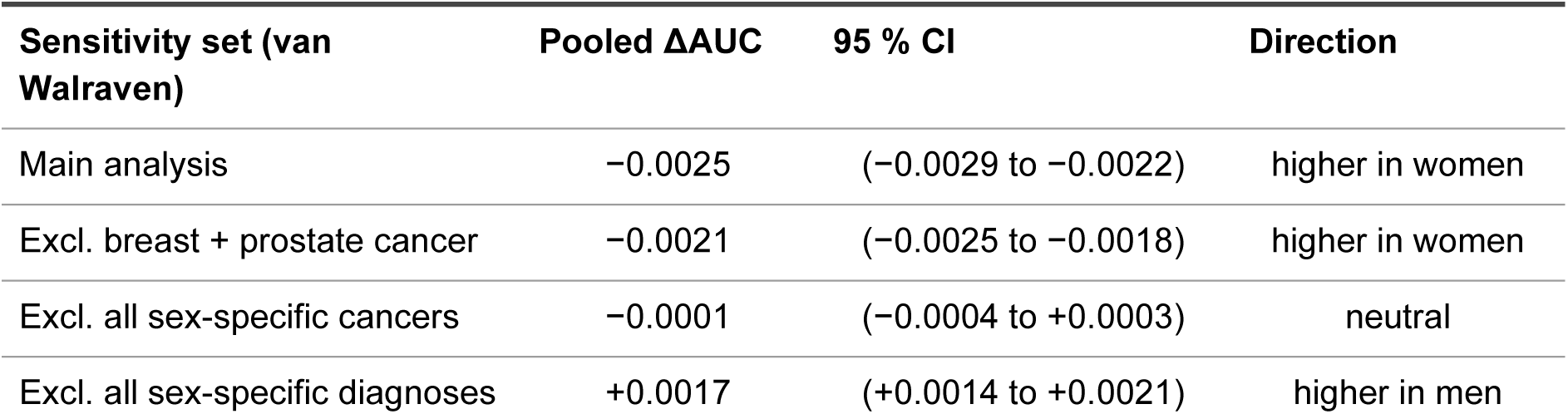
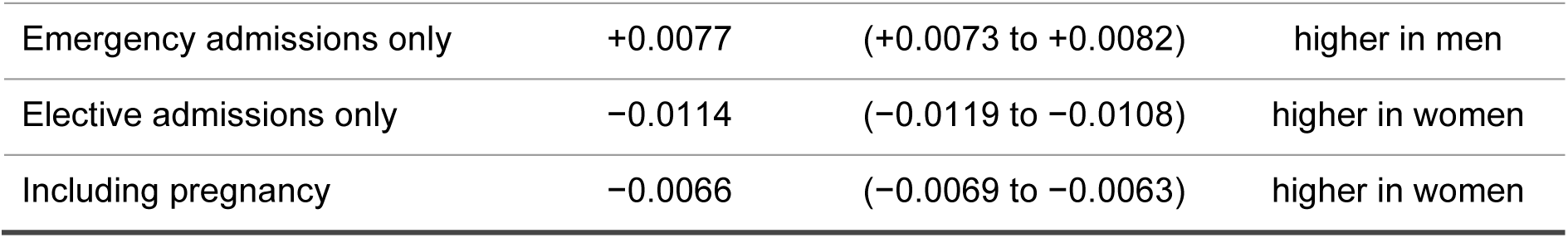
Sensitivity analyses for the van Walraven score, meta-pooled. Sets were pre-specified in the OSF pre-registration. Sensitivity sets and the main-analysis reference row are pooled by fixed-effects inverse-variance meta-analysis, consistent with Figure 6; the corresponding random-effects main-analysis estimate is −0.0026 (Table 5). The reversal between emergency and elective admissions motivates the separate clinical-utility analysis.

| Sensitivity set (van Walraven) | Pooled $\Delta AUC$ | 95 % CI | Direction |
| --- | --- | --- | --- |
| Main analysis | $-0.0025$ | $(-0.0029 \text{ to } -0.0022)$ | higher in women |
| Excl. breast + prostate cancer | $-0.0021$ | $(-0.0025 \text{ to } -0.0018)$ | higher in women |
| Excl. all sex-specific cancers | $-0.0001$ | $(-0.0004 \text{ to } +0.0003)$ | neutral |
| Excl. all sex-specific diagnoses | $+0.0017$ | $(+0.0014 \text{ to } +0.0021)$ | higher in men |

**Table 6.** Sensitivity analyses for the van Walraven score, meta-pooled.
| Sensitivity set (van Walraven) | Pooled ΔAUC | 95 % CI | Direction |
| --- | --- | --- | --- |
| Emergency admissions only | +0.0077 | (+0.0073 to +0.0082) | higher in men |
| Elective admissions only | -0.0114 | (-0.0119 to -0.0108) | higher in women |
| Including pregnancy | -0.0066 | (-0.0069 to -0.0063) | higher in women |

### Age-stratified analysis and COVID-era comparison

The sex-related difference is consistent across age strata 18 to 79 years and attenuated in the ≥ 80 years stratum (Table 7, Figure 7). The Elixhauser-Sum ΔAUC reverses direction in the oldest stratum. The absolute discrimination of all three scores decreases steeply with age, from c-statistics of approximately 0.82 to 0.95 in those aged 18 to 44 years (depending on the score) to approximately 0.63 to 0.71 in those aged ≥ 80 years. Four mechanisms plausibly contribute to this age-dependence. First, the baseline mortality rises with age (from approximately 0.4 % in those under 45 to over 12 % in those aged ≥ 80), and at high event prevalence the score has less room to separate high-risk from low-risk patients. Second, age itself becomes a dominant mortality driver at advanced age, and the relative predictive contribution of additional comorbidities diminishes. Third, survival selection homogenises the very-elderly hospitalised population, since patients with severe pre-existing conditions are less likely to reach hospital at age ≥ 80 alive. Fourth, multidimensional frailty, which discharge-coded comorbidity domains do not fully capture, becomes the dominant mortality driver above age 80; this is the central observation of Frenkel and colleagues in acute-geriatric patients [28] and is consistent with the long-term-mortality discrimination decay reported by Westerberg and colleagues [17]. Because age is only a crude proxy for menopausal status, these strata cannot test whether hormonal factors contribute to the sex-related difference, and we do not interpret the age pattern in hormonal terms. Across the COVID era (2020 to 2023) all three scores show a small absolute decrease in discrimination compared with pre-COVID (2010 to 2019), slightly larger in men than in women (Table 8). The sign of the sex-related difference persists throughout. In reporting year 2024, in-hospital mortality has only partially returned to pre-pandemic levels (Figure 8) and the score-discrimination values are close to the 2020 to 2023 mean.

**Figure 7.**
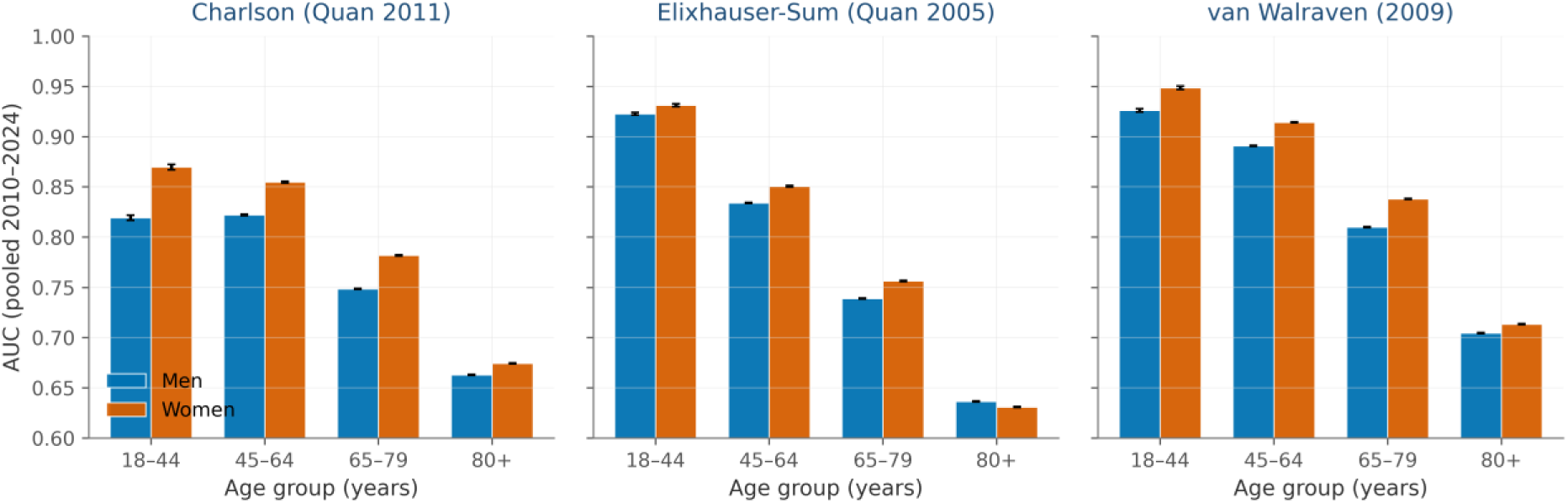
Discrimination by age group, sex, and score (pooled).

**Figure 8.**
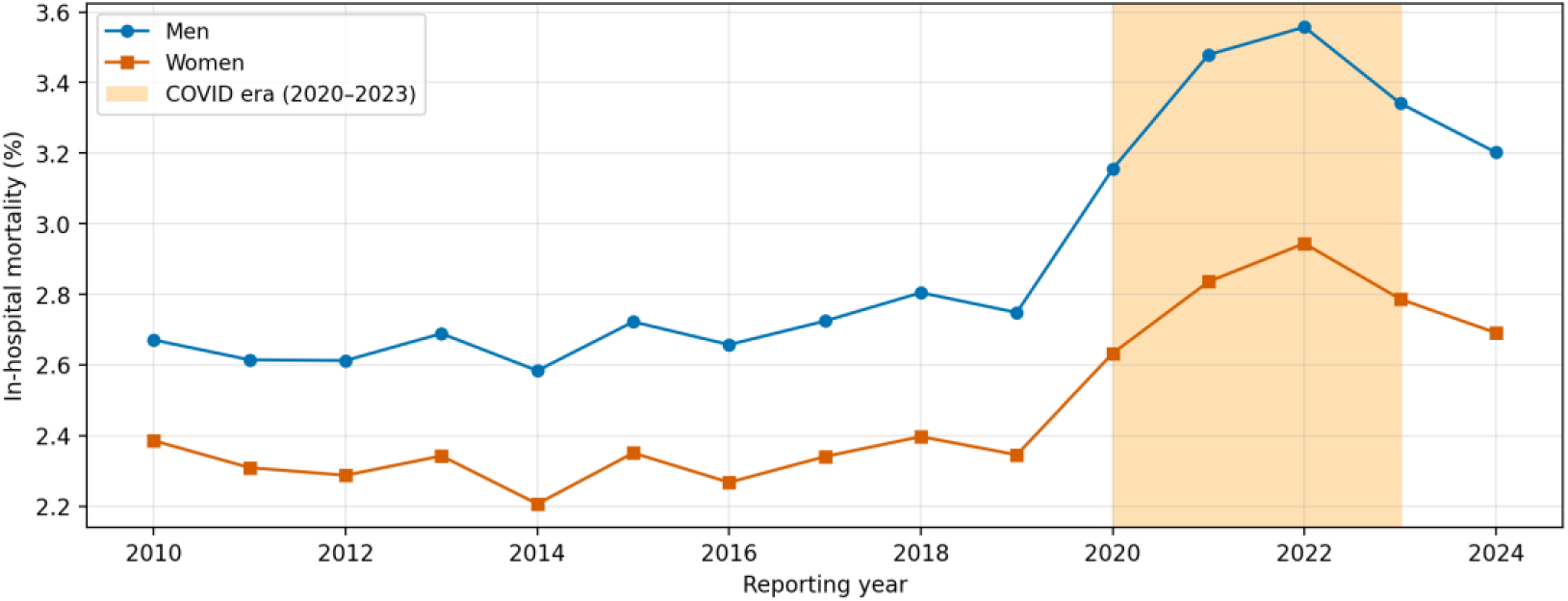
In-hospital mortality by reporting year and sex. COVID era (2020 to 2023) shaded; reporting year 2024 is the post-emergency phase and has only partially returned to pre-pandemic levels.

**Table 7.** Age-stratified pooled discrimination by score and sex, pooled 2010–2024.

| Score | Age (yrs) | AUC men / women | ΔAUC (95 % CI) | z |
| --- | --- | --- | --- | --- |
| Charlson | 18–44 | 0.819 / 0.870 | –0.051 (–0.054 to –0.047) | –26.9 |
|  | 45–64 | 0.822 / 0.855 | –0.033 (–0.034 to –0.032) | –60.3 |
|  | 65–79 | 0.748 / 0.782 | –0.034 (–0.034 to –0.033) | –89.5 |
| | $\geq 80$ | 0.663 / 0.674 | –0.011 (–0.012 to –0.011) | –32.3 |
| Elixhauser-Sum | 18–44 | 0.923 / 0.931 | –0.009 (–0.011 to –0.007) | –8.8 |
|  | 45–64 | 0.834 / 0.851 | –0.017 (–0.018 to –0.016) | –36.6 |
|  | 65–79 | 0.739 / 0.756 | –0.018 (–0.018 to –0.017) | –45.8 |
| | $\geq 80$ | 0.636 / 0.631 | +0.005 (+0.005 to +0.006) | +14.3 |
| van Walraven | 18–44 | 0.926 / 0.949 | –0.022 (–0.025 to –0.020) | –19.2 |
|  | 45–64 | 0.891 / 0.914 | –0.023 (–0.024 to –0.023) | –63.8 |
|  | 65–79 | 0.810 / 0.838 | –0.028 (–0.029 to –0.028) | –90.1 |
| | $\geq 80$ | 0.704 / 0.713 | –0.009 (–0.009 to –0.008) | –26.2 |

**Table 8.** Pre-COVID (2010 to 2019) versus COVID era (2020 to 2023) discrimination by sex. Pre-COVID and COVID-era values are means of the per-year AUC across the respective reporting years.

| Score | Sex | Pre-COVID AUC (2010 to 2019) | COVID-era AUC (2020 to 2023) | $\Delta$ AUC (COVID – pre) |
| --- | --- | --- | --- | --- |
| Charlson | Men | 0.784 | 0.752 | –0.032 |
|  | Women | 0.790 | 0.765 | –0.025 |
| Elixhauser-Sum | Men | 0.782 | 0.766 | –0.017 |
|  | Women | 0.773 | 0.761 | –0.012 |
| van Walraven | Men | 0.837 | 0.817 | –0.020 |
|  | Women | 0.839 | 0.822 | –0.017 |

## Discussion

In a complete national enumeration of 166 693 491 German adult acute somatic inpatient cases from 2010 to 2024, the three most widely used comorbidity scores in administrative-data risk adjustment performed differently in women and men, and the difference lay in calibration rather than in ranking. Calibration-in-the-large was effectively perfect for both sexes, so the scores were correct on average. What differed was the calibration slope, meaning that predicted risk tracked observed risk at a different rate in women than in men, and in opposite directions. At the same score, men died more often than women. Discrimination differed only marginally. The direction was consistent across all 15 reporting years and across the age range, with a single exception among the oldest patients, and it persisted in four of the seven analysis sets. It attenuated to neutrality when all sex-specific cancers were excluded, reversed when all sex-specific diagnoses were excluded, and reversed between emergency and elective admissions, which points to case-mix as a substantial part of the mechanism.

Several limitations bound what these findings can support. Comorbidities are identified from discharge diagnosis codes, so they reflect what was documented and coded for reimbursement rather than a clinical assessment of severity, and chronic conditions may be under-coded at an acute admission. German Diagnosis-Related Group data carry no present-on-admission flag, so pre-existing conditions cannot be separated reliably from complications arising during the stay, which introduces partial reverse causation. Because all three scores share this feature of the data source, the comparison between scores is unaffected, but the absolute calibration of any single score is not. Comorbidities are taken from the index admission alone, without a longitudinal look-back, which is known to weaken comorbidity adjustment [29]. Sex is available only as the administrative binary recorded in the statistics: Cases registered under the third German legal category are too few to analyse separately and are not identifiable in the aggregated outputs, and no variable captures gender as a social dimension, so nothing in this study speaks to gender identity or to gender-related care pathways. Standard errors are not adjusted for clustering within hospitals, because hospital identifiers are not part of the aggregated outputs, so the confidence intervals are somewhat narrower than warranted [30]. The outcome is in-hospital death, which depends on discharge practice. The analysis plan was fixed in the time-stamped Research Data Centre submission syntax before any aggregated output existed, but the Open Science Framework entry was filed after the outputs had been returned and quality-checked and is declared as retrospective there [20], so the study has no prospectively registered public plan and readers should weigh the syntax provenance (Supplementary File S2) accordingly. Generalisability is bounded by the setting: The cohort covers acute somatic care in Germany and excludes psychiatric admissions billed under a separate system (PEPP), which are female-predominant, as well as rehabilitation admissions, so the magnitude reported here should not be transferred to other health systems without re-validation. These and four further considerations, including the linearity assumption of the calibration slope, end-of-life coding and the choice of in-hospital rather than 30-day mortality, are set out in full in Supplementary File S3. Set against this, the study uses a census rather than a sample over 15 consecutive years, evaluates all three scores in parallel on identical records with an identical fitting procedure, pre-specified calibration as the primary axis and judged effects against pre-specified effect-size thresholds rather than p-values, complements the model-based slope with a non-parametric comparison of observed mortality per score quintile, verified the pooling pipeline along three independent calculation paths, and reports all results disaggregated by sex in line with SAGER 2016 [4].

These calibration findings replicate and extend earlier population-based work. Rius and colleagues reported good Charlson calibration in men but poor calibration in women in a Catalan cohort [16], a pattern consistent with a difference located on the calibration axis. Westerberg and colleagues showed in a Swedish nationwide cohort that the Charlson index discriminates long-term mortality well and that discrimination improves with a longer look-back [17], which is relevant to our single-admission limitation, although both the outcome and the time horizon differ from ours. Neither study evaluated all three scores, and neither addressed in-hospital mortality at national scale. That the direction of the slope deviation is opposite between the Charlson index and the Elixhauser-Sum, although both were computed from identical records with an identical fitting procedure, locates the difference in how each score selects and weights comorbidity domains rather than in the underlying data; the developmental heterogeneity of the three constructions is discussed in Supplementary File S3.

A plausible explanation for the direction of the difference lies in how administrative scores encode illness. Each score registers a condition as a binary code, not its severity, yet both the severity behind a given code and the propensity to document it differ between women and men. Men in this cohort carried more of the conditions most tightly linked to death, including mechanical ventilation, liver disease, metastatic cancer and cardiac arrhythmia (Table 1), so at an identical score their underlying illness is on average more severe, which is consistent with their higher observed mortality at the same score. Conditions that are documentation-sensitive and more often coded in women, such as depression, can raise a score without a matching rise in mortality risk. Sex therefore acts as a modifier of the relationship between score and outcome in the sense set out by Mauvais-Jarvis and colleagues [1], and a single calibration applied to both sexes cannot absorb it. For an individual patient the difference is unlikely to change a clinical decision: A slope difference of this size shifts absolute risk by roughly 0.2 percentage points at the cohort mean mortality of 2.7 % and by about 1 percentage point in the highest-risk quintile. The consequence lies elsewhere. These scores are reused at scale, entering a large body of observational studies, national quality-reporting systems and, increasingly, clinical decision-support algorithms, almost always with one formula for both sexes, as in nationwide German outcome research [31] and in the national hospital quality-measurement framework [32]. A small but systematic and directionally stable difference does not average out under reuse but is reproduced in every SMR and every case-mix adjustment that ingests the score. Published comparisons of risk-adjustment methods give a sense of the plausible scale: Adding present-on-admission indicators shifted 21 % of hospital rankings by at least two deciles [33], altered statistical methodology moved 14 % of surgeon rankings [34], and switching between in-hospital and 30-day outcomes moved 10 to 15 % of hospital quintile classifications [35]. As an illustration rather than a measured effect, two hospitals with identical comorbidity case-mix but a 30-percentage-point difference in the proportion of men would differ in expected-versus-observed alignment by roughly 4 % from sex composition alone (0.30 × the 13 % contrast between an all-male and an all-female case-mix at matched Charlson score), and model miscalibration is an established driver of such benchmarking error [36]. We do not claim to have measured the downstream effect, which requires patient-to-hospital linkage; we do claim that dismissing it as negligible is unwarranted, because its magnitude is unquantified rather than known to be small. The difference can be addressed either by recalibrating the scores separately for women and men or by adding sex to the risk-adjustment model.

Under the equity requirements of the EU AI Act 2024 [7], whether retrospective risk-adjustment scores used for hospital benchmarking fall within scope is not yet settled; where such scores inform high-risk decision support, an equity assessment would have to cover calibration and not only discrimination, because calibration is the axis on which these scores differ between the sexes. The concerns about algorithmic equity raised by Obermeyer [37] and Rajkomar [38] apply equally to non-machine-learning instruments such as these three scores, and sex and gender analysis has been argued to strengthen the rigour and reproducibility of research precisely because designs that ignore sex allow systematic error to propagate unchecked [39]. Three questions follow. First, whether recalibrating the three scores with sex-specific weights removes the difference, using the Swiss [40] and AHRQ [14] recalibration templates as a model. Second, how large the consequences for hospital-comparison metrics actually are, which requires patient-to-hospital linkage, present-on-admission indicators and a 30-day outcome. Third, whether the reversal between emergency and elective admissions is explained by case-mix, which an ICD-chapter-stratified analysis of the same data would test.

## Conclusion

Applied with a single formula to both sexes, three established comorbidity scores are calibrated differently in women and men in the German adult inpatient population. The difference is largest for the Charlson index, while discrimination differs only marginally. Because these instruments are used with one formula for both sexes across a large body of research and quality reporting, this difference in calibration is the consequential result: small for the individual patient, but systematic, stable in direction and reproduced wherever the scores are reused. It supports investigating sex-specific recalibration of established risk-adjustment instruments; the full consequences for hospital benchmarking remain to be quantified rather than assumed small.

## Supporting information

RECORD Checklist

SAGER Checklist

STROBE Checklist

Supplementary File S1: Comorbidity definitions and score weights

Supplementary File S2: Pre-specified analysis code submitted to the Research Data Centre

Supplementary File S3: Extended methodological considerations

TRIPOD+AI Checklist

## Acknowledgements

We thank the Research Data Centre (FDZ) of the Federal Statistical Office and the Statistical Offices of the Federal States for access to the DRG statistics under Project 4509-2021 [21] and for execution of the analysis syntax.

## Supplementary material

S1, comorbidity definitions and score component weights; S2, the time-stamped Research Data Centre submission syntax; S3, extended methodological considerations. Reporting checklists (STROBE, RECORD, TRIPOD+AI, SAGER) are provided as submission appendices.

## Data availability statement

Aggregated output files and analysis code will be deposited on the Open Science Framework (DOI 10.17605/OSF.IO/P3QAW) upon publication. The underlying individual-level data are held by the Research Data Centre of the German Federal Statistical Office and are not transferable due to national statistics confidentiality rules (§16 BStatG). Replication is possible by separate research-project application to the FDZ under the same Controlled Remote Data Processing access route. The data product used is cited as reference 21.

## License

This preprint is made available under a Creative Commons Attribution 4.0 International License (CC BY 4.0).

