## Supplementary material for "Do established comorbidity scores predict in-hospital mortality equally well in women and men? A nationwide validation in 166.7 million German inpatient cases, 2010–2024": RECORD Checklist

### RECORD Reporting Checklist

Reproduces the RECORD-specific items of the checklist that extends STROBE for studies using routinely collected health data (Benchimol EI, Smeeth L, Guttmann A, Harron K, Moher D, Petersen I, Sørensen HT, von Elm E, Langan SM; RECORD Working Committee. PLoS Med 2015;12(10):e1001885; checklist under CC BY). The STROBE items themselves are addressed in the separate STROBE checklist.

| Item | RECORD item | Location and compliance |
|---|---|---|
| 1.1 | The type of data used should be specified in the title or abstract; where possible the name of the databases used should be included. | Title names German inpatient cases; Abstract (Database, Setting) names the German Diagnosis-Related Group (DRG) statistics under §21 of the Hospital Reimbursement Act, accessed through the Research Data Centre of the Federal Statistical Office. |
| 1.2 | If applicable, the geographic region and timeframe should be reported in the title or abstract. | Title states Germany and 2010–2024; Abstract (Setting) repeats country and reporting years. |
| 1.3 | If linkage between databases was conducted, this should be clearly stated in the title or abstract. | No linkage was performed. The analysis uses a single data source; this is stated in Methods (Data source and cohort construction) and in the Data availability statement. |
| 6.1 | The methods of study population selection, such as codes or algorithms used to identify subjects, should be listed in detail. | Methods (Data source and cohort construction) lists the nine eligibility steps in fixed order with the operational definitions: age 18 to 110 years; admission-reason codes 1 and 2; absence of admission-origin codes V, A, K; absence of discharge codes 6, 8, 13, 29; length of stay at least 2 days; sex documented; principal diagnosis not in ICD-10 chapter O; principal diagnosis not ICD-10-GM Z51; no accounting-only department codes. |
| 6.2 | Any validation studies of the codes or algorithms used to select the population should be referenced; if validation was conducted for this study, methods and results should be provided. | The comorbidity algorithms are the published and independently validated mappings of Quan and colleagues 2005 and 2011 and van Walraven and colleagues 2009, cited in Methods; the exact ICD-10-GM implementation is reproduced in Supplementary File S1. The eligibility algorithm itself has not been separately validated against clinical records, which is stated as a limitation. |
| 6.3 | If the study involved linkage of databases, consider a flow diagram or other graphical display of the linkage process. | Not applicable, no linkage. A cohort attrition flow diagram covering all nine eligibility steps and all 15 reporting years is provided as Figure 1. |
| 7.1 | A complete list of codes and algorithms used to classify exposures, outcomes, confounders and effect modifiers should be provided. | Supplementary File S1 gives the complete ICD-10-GM code definitions for all 31 Elixhauser domains and all 17 Charlson conditions together with the van Walraven point weights and the Charlson integer weights. The outcome is the administrative discharge code for death, defined in Methods (Outcome and predictors). |
| 12.1 | Authors should describe the extent to which the investigators had access to the database population used to create the study population. | Methods (Study design and registration; Data source and cohort construction) state that access is by controlled remote data processing: the authors never accessed individual-level records, the analysis syntax was executed by Research Data Centre staff on the original data, and only aggregated output subject to a minimum cell size left the Centre. |
| 12.2 | Authors should provide information on the data cleaning methods used in the study. | Because individual records were not accessible, no case-level cleaning was possible or performed. Quality control of the returned aggregated outputs comprised summation checks against the cohort flow diagram, range plausibility checks of |

| | | |
|---|---|---|
| | | the performance metrics, and per-set row-count checks; this is described in Methods (Verification, software, reporting standards, ethics, patient and public involvement). |
| 12.3 | State whether the study included person-level, institutional-level or other data linkage across two or more databases, with methods of linkage and of linkage-quality evaluation. | No linkage of any kind was performed. The absence of patient-to-hospital linkage is named in the Discussion as the reason why the downstream benchmarking effect could not be quantified, and the absence of cross-admission patient linkage is named as the reason for the single-admission look-back. |
| 13.1 | Describe in detail the selection of persons included in the study, including filtering based on data quality, data availability and linkage. | Figure 1 and Methods report the per-step exclusion counts from 266 046 985 raw admissions to the final 166 693 491 cases, together with the year-to-year variability of each exclusion share and the rising trend of the short-stay step from 12.9 % in 2010 to 17.4 % in 2024. |
| 19.1 | Discuss the implications of using data that were not created or collected to answer the specific research question, including misclassification bias, unmeasured confounding, missing data and changing eligibility over time. | Strengths and limitations addresses this directly: comorbidities reflect what was documented and coded for reimbursement rather than clinical severity, chronic conditions may be under-coded at an acute admission, no present-on-admission flag permits separation of pre-existing conditions from complications arising during the stay, absence of an ICD code is interpreted as absence of the condition, and coding and eligibility patterns drift over the 15 years. The Discussion sections on present-on-admission coding and on documentation versus clinical effects develop the misclassification argument in detail. |
| 22.1 | Authors should provide information on how to access supplemental information such as the study protocol, raw data or programming code. | The Data availability statement gives the Open Science Framework deposition (DOI 10.17605/OSF.IO/P3QAW) for aggregated outputs and analysis code, states that individual-level data are not transferable under §16 of the Federal Statistics Act, and describes the separate application route for independent replication. The time-stamped Research Data Centre submission syntax is reproduced in full as Supplementary File S2. |
