## Supplementary material for "Do established comorbidity scores predict in-hospital mortality equally well in women and men? A nationwide validation in 166.7 million German inpatient cases, 2010–2024": SAGER Checklist

### SAGER Reporting Checklist — Sex and Gender Equity in Research

*Filled for: Sex-differential calibration of three established comorbidity scores applied to in-hospital mortality (German DRG statistics 2010–2024)*

Reproduces the SAGER guideline recommendations (Heidari S, Babor TF, De Castro P, Tort S, Curno M. Sex and Gender Equity in Research: rationale for the SAGER guidelines and recommended use. Res Integr Peer Rev 2016;1:2). Terminology: the term "sex" (biological, administrative-binary as recorded in the DRG statistics) is used throughout; "gender" is not analysed and this is stated explicitly.

| SAGER area | Recommendation | Location in manuscript and compliance |
|---|---|---|
| Title and abstract | State that the study concerns both sexes and that analyses are sex-disaggregated; use the terms sex and gender correctly. | Title poses the sex-comparison question directly; Abstract (Objectives, Results, Conclusions) reports the sex-stratified analysis for men and women. "Sex" is used throughout, not "gender". |
| Introduction | Report whether sex and/or gender differences may be expected and the rationale for examining them. | The Introduction opens with the sex and gender equity framing, citing the systematic under-consideration of sex and gender in clinical research and the move to sex-specific risk models, and defines sex and gender explicitly at first mention: sex is used in the biological and administrative sense recorded in the data, gender as the sociocultural dimension, with the explicit statement that gender is not represented in the dataset and that no inference about gender identity or gender-related care pathways is drawn. |
| Methods | Report how sex was accounted for in the design, how it was determined, and how data were disaggregated; define the variable. | Methods (Outcome and predictors; Statistical analysis) specify sex as the administrative-binary variable recorded in the DRG statistics, a sex-blind model evaluated in sex-stratified subgroups, and state explicitly that "sex" (not "gender") is used per SAGER 2016 [19]. |

| | | |
|---|---|---|
| Results | Present data disaggregated by sex. | All primary and secondary results are reported by sex: cohort characteristics (Table 1), calibration (Table 2, Figure 2), observed mortality per score quintile (Table 3), discrimination (Tables 4–5), sensitivity, age strata and COVID era (Tables 6–8). |
| Discussion | Discuss the implications of sex on the findings and note limitations where sex was not fully captured. | Discussion interprets the sex-differential calibration and its downstream relevance; limitations state that sex is recorded as administrative-binary, with no representation of non-binary categories or gender identity, and that age is only a crude proxy for menopausal status. |
