## Supplementary material for "Do established comorbidity scores predict in-hospital mortality equally well in women and men? A nationwide validation in 166.7 million German inpatient cases, 2010–2024": STROBE Checklist

### STROBE Statement — Checklist for cohort studies

Filled for: Sex-asymmetric calibration of three established comorbidity scores applied to in-hospital mortality (German DRG statistics 2010–2024)

*Reproduces the STROBE v4 checklist for cohort, case-control, and cross-sectional studies (cohort column), www.strobe-statement.org. Section references point to manuscript HERA_Paper1_v1.8; final page/line numbers to be inserted from the typeset proof. Submit as a separate reporting-checklist document (not a numbered scientific supplement).*

| Item | Recommendation | Reported in (manuscript section) |
|---|---|---|
| **§ Title and abstract** | | |
| 1 (a) | Indicate the study's design with a commonly used term in the title or the abstract | Title; Abstract — Design |
| 1 (b) | Provide in the abstract an informative and balanced summary of what was done and what was found | Abstract; Plain Language Summary |
| **§ Introduction** | | |
| 2 | Background/rationale: explain the scientific background and rationale for the investigation being reported | Introduction |
| 3 | Objectives: state specific objectives, including any prespecified hypotheses | Introduction (final paragraph, hypotheses); Abstract — Objectives |
| **§ Methods** | | |
| 4 | Study design: present key elements of study design early in the paper | Methods — Study design and registration; Abstract — Design |
| 5 | Setting: describe the setting, locations, and relevant dates, including periods of recruitment, exposure, follow-up, and data collection | Methods — Data source and cohort construction (DRG statistics 2010–2024); Abstract — Setting |
| 6 (a) | Participants: give the eligibility criteria, and the sources and methods of selection of participants; describe methods of follow-up | Methods — Data source and cohort construction (nine-step eligibility filter); Figure 1 |
| 6 (b) | For matched studies, give matching criteria and number of exposed and unexposed | Not applicable (no matching; full enumeration, sex-stratified) |
| 7 | Variables: clearly define all outcomes, exposures, predictors, potential confounders, and effect modifiers; give diagnostic criteria, if applicable | Methods — Outcome and predictors; Supplementary File S1 (comorbidity definitions) |
| 8 | Data sources/measurement: for each variable of interest, give sources of data and details of methods of assessment (measurement); describe comparability of assessment methods if there is more than one group | Methods — Data source and cohort construction; Outcome and predictors; Supplementary File S1 |
| 9 | Bias: describe any efforts to address potential sources of bias | Methods — Statistical analysis and fairness assessment; Discussion — Other methodological considerations; Strengths and limitations |
| 10 | Study size: explain how the study size was arrived at | Methods — Data source (complete enumeration; n = 166 693 491); Abstract — Participants |
| 11 | Quantitative variables: explain how quantitative variables were handled in the analyses; if applicable, describe which groupings were chosen and why | Methods — Outcome and predictors (score construction); Statistical analysis (score quintiles) |
| 12 (a) | Statistical methods: describe all statistical methods, including those used to control for confounding | Methods — Statistical analysis and fairness assessment |
| 12 (b) | Describe any methods used to examine subgroups and interactions | Methods — Statistical analysis (sex-stratified; age strata; admission type) |
| 12 (c) | Explain how missing data were addressed | Methods — Data source (sex-documented inclusion step); Statistical analysis |
| 12 (d) | Cohort study — if applicable, explain how loss to follow-up was addressed | Not applicable (in-hospital outcome at discharge; no post-discharge follow-up) |
| 12 (e) | Describe any sensitivity analyses | Methods — Statistical analysis; Results — Sensitivity analyses (seven sets); Pre-specified clinical-meaningfulness thresholds |
| **§ Results** | | |

| | | |
|---|---|---|
| 13 (a) | Participants: report numbers of individuals at each stage of study (eg, eligible, examined, confirmed eligible, included, analysed) | Results — Cohort characteristics; Figure 1 (cohort flow diagram); Methods — Data source |
| 13 (b) | Give reasons for non-participation at each stage | Methods — Data source (per-step exclusion shares); Figure 1 |
| 13 (c) | Consider use of a flow diagram | Figure 1 (cohort flow diagram) |
| 14 (a) | Descriptive data: give characteristics of study participants and information on exposures and potential confounders | Results — Cohort characteristics; Table 1 |
| 14 (b) | Indicate number of participants with missing data for each variable of interest | Methods — Data source; Results — Cohort characteristics (sex-documented cohort) |
| 14 (c) | Cohort study — summarise follow-up time | Not applicable (in-hospital outcome; length of stay reported in Table 1) |
| 15 | Outcome data: cohort study — report numbers of outcome events or summary measures over time | Results — Cohort characteristics; Table 1; Figure 8 (mortality by year and sex) |
| 16 (a) | Main results: give unadjusted estimates and, if applicable, confounder-adjusted estimates and their precision (eg, 95% CI) | Results — Calibration by sex; Discrimination by sex; Tables 2–5, 7, 8 |
| 16 (b) | Report category boundaries when continuous variables were categorized | Results — Observed mortality per score quintile; Table 3 (quintile cut-points) |
| 16 (c) | If relevant, consider translating estimates of relative risk into absolute risk for a meaningful time period | Results — Observed mortality per score quintile (absolute mortality); Discussion — Implications (absolute risk shift) |
| 17 | Other analyses: report other analyses done — eg analyses of subgroups and interactions, and sensitivity analyses | Results — Sensitivity analyses; Age-stratified analysis and COVID-era comparison; Tables 6–8 |
| **§ Discussion** | | |
| 18 | Key results: summarise key results with reference to study objectives | Discussion (opening paragraph); Conclusion |
| 19 | Limitations: discuss limitations of the study, taking into account sources of potential bias or imprecision; discuss both direction and magnitude of any potential bias | Discussion — Strengths and limitations; Other methodological considerations |
| 20 | Interpretation: give a cautious overall interpretation of results considering objectives, limitations, multiplicity of analyses, results from similar studies, and other relevant evidence | Discussion — Interpretation and the subsequent subsections; Implications and research priorities |
| 21 | Generalisability: discuss the generalisability (external validity) of the study results | Discussion — Strengths and limitations; Implications and research priorities |
| **§ Other information** | | |
| 22 | Funding: give the source of funding and the role of the funders for the present study and, if applicable, for the original study on which the present article is based | Funding (None.) |
