## Supplementary File S1: Comorbidity definitions and score weights for "Do established comorbidity scores predict in-hospital mortality equally well in women and men? A nationwide validation in 166.7 million German inpatient cases, 2010–2024"

### Comorbidity definitions and score weights

*Supplement to: Sex-asymmetric calibration of three established comorbidity scores applied to in-hospital mortality. German DRG statistics 2010 to 2024.*

*Study registration OSF DOI 10.17605/OSF.IO/P3QAW*

### How the scores were computed

Comorbidities were identified from the principal discharge diagnosis and all secondary discharge diagnoses of each case, combined into one diagnosis string per admission. A comorbidity domain was flagged when any of its listed ICD-10-GM code stems appeared in that string. Code definitions follow the enhanced ICD-10 coding algorithm of Quan and colleagues (2005), applied to the German modification ICD-10-GM.

Three scores were derived per case from these domain flags. The Elixhauser Comorbidity Sum is the unweighted count of the 31 domains in Table S1.1, each present domain counting as one. The van Walraven score is the weighted sum of the 21 domains that carry a point value in Table S1.1, using the point system of van Walraven and colleagues (2009). The Charlson Comorbidity Index uses the 17 conditions and integer weights of Quan and colleagues (2011), listed separately in Table S1.2, because several Charlson phenotypes are defined differently from the Elixhauser domains. Scores were computed in Stata 19; the exact computation code is provided in Supplementary File S3.

**Table S1.1.** *Elixhauser comorbidity domains (ICD-10-GM code definitions per Quan 2005) and van Walraven point weights.*

| Comorbidity domain | ICD-10-GM code stems | van Walraven points |
|---|---|---|
| Cardiac arrhythmias | I47, I48, I49, I441, I442, I443, I456, I459, R000, R001, R008, T821, Z450, Z950 | +5 |
| Valvular disease | I05, I06, I07, I08, I34, I35, I36, I37, I38, I39, I091, I098, A520, Q230, Q231, Q232, Q233, Z952, Z953, Z954 | -1 |
| Pulmonary circulation disorders | I26, I27, I280, I288, I289 | +4 |
| Peripheral vascular disorders | I70, I71, I731, I738, I739, I771, I790, I792, K551, K558, K559, Z958, Z959 | +2 |
| Hypertension, uncomplicated | I10 | — |
| Hypertension, complicated | I11, I12, I13, I15 | — |
| Congestive heart failure | I43, I50, I099, I110, I130, I132, I255, I420, I425, I426, I427, I428, I429, P290 | +7 |
| Paralysis | G81, G82, G041, G114, G801, G802, G830, G831, G832, G833, G834, G839 | +7 |

| Comorbidity domain | ICD-10-GM code stems | van Walraven points |
|---|---|---|
| Other neurological disorders | G10, G11, G12, G13, G20, G21, G22, G32, G35, G36, G37, G40, G41, G254, G255, G312, G318, G319, G931, G934, R470, R56 | +6 |
| Chronic pulmonary disease | J40, J41, J42, J43, J44, J45, J46, J47, J60, J61, J62, J63, J64, J65, J66, J67, I278, I279, J684, J701, J703 | +3 |
| Diabetes, uncomplicated | E100, E101, E109, E110, E111, E119, E120, E121, E129, E130, E131, E139, E140, E141, E149 | — |
| Diabetes, complicated | E102, E103, E104, E105, E106, E107, E108, E112, E113, E114, E115, E116, E117, E118, E122, E123, E124, E125, E126, E127, E128, E132, E133, E134, E135, E136, E137, E138, E142, E143, E144, E145, E146, E147, E148 | — |
| Hypothyroidism | E00, E01, E02, E03, E890 | — |
| Renal failure | N18, N19, I120, I131, N250, Z490, Z491, Z492, Z940, Z992 | +5 |
| Liver disease | B18, I85, K70, K72, K73, K74, I864, I982, K711, K713, K714, K715, K717, K760, K762, K763, K764, K765, K766, K767, K768, K769, Z944 | +11 |
| Peptic ulcer disease (excluding bleeding) | K257, K259, K267, K269, K277, K279, K287, K289 | — |
| AIDS / HIV | B20, B21, B22, B24 | — |
| Lymphoma | C81, C82, C83, C84, C85, C88, C96, C900, C902 | +9 |
| Metastatic cancer | C77, C78, C79, C80 | +12 |
| Solid tumour without metastasis | C00, C01, C02, C03, C04, C05, C06, C07, C08, C09, C10, C11, C12, C13, C14, C15, C16, C17, C18, C19, C20, C21, C22, C23, C24, C25, C26, C30, C31, C32, C33, C34, C37, C38, C39, C40, C41, C43, C45, C46, C47, C48, C49, C50, C51, C52, C53, C54, C55, C56, C57, C58, C59, C60, C61, C62, C63, C64, C65, C66, C67, C68, C69, C70, C71, C72, C73, C74, C75, C76, C97 | +4 |
| Rheumatoid arthritis / collagen vascular disease | L940, L941, L943, M05, M06, M08, M30, M32, M33, M34, M35, M45, M120, M123, M310, M311, M312, M313, M461, M468, M469 | — |
| Coagulopathy | D65, D66, D67, D68, D691, D693, D694, D695, D696 | +3 |
| Obesity | E66 | -4 |
| Weight loss | E40, E41, E42, E43, E44, E45, E46, R634, R64 | +6 |
| Fluid and electrolyte disorders | E86, E87, E222 | +5 |
| Blood loss anaemia | D500 | -2 |
| Deficiency anaemia | D51, D52, D53, D508, D509 | -2 |
| Alcohol abuse | E52, F10, T51, G621, I426, K292, K700, K703, K709, Z502, Z714, Z721 | — |
| Drug abuse | F11, F12, F13, F14, F15, F16, F18, F19, Z715, Z722 | -7 |
| Psychoses | F20, F22, F23, F24, F25, F28, F29, F302, F312, F315 | — |
| Depression | F32, F33, F204, F313, F314, F315, F341, F412, F432 | -3 |

A domain is flagged when any listed code stem appears among the principal or secondary diagnoses. The van Walraven points column applies to the van Walraven score only; domains marked — contribute to the Elixhauser Comorbidity Sum but not to the van Walraven score. The Elixhauser Comorbidity Sum counts each domain present as one, without weighting. Hypertension is split into an uncomplicated and a complicated domain. Code definitions follow Quan et al., Med Care 2005;43:1130–9; van Walraven point weights follow van Walraven et al., Med Care 2009;47:626–33.

**Table S1.2.** *Charlson Comorbidity Index conditions and integer weights (Quan 2011 update).*

| Condition | ICD-10-GM code stems | Charlson weight |
|---|---|---|
| Myocardial infarction | I21, I22, I252 | 0 |
| Congestive heart failure | I43, I50, I099, I110, I130, I132, I255, I420, I425, I426, I427, I428, I429, P290 | 2 |
| Peripheral vascular disease | I70, I71, I731, I738, I739, I771, I790, I792, K551, K558, K559, Z958, Z959 | 0 |
| Cerebrovascular disease | G45, G46, H340, I60, I61, I62, I63, I64, I65, I66, I67, I68, I69 | 0 |
| Dementia | F00, F01, F02, F03, F051, G30, G311 | 2 |
| Chronic pulmonary disease | J40, J41, J42, J43, J44, J45, J46, J47, J60, J61, J62, J63, J64, J65, J66, J67, I278, I279, J684, J701, J703 | 1 |
| Rheumatic disease | M05, M06, M315, M32, M33, M34, M351, M353, M360 | 1 |
| Peptic ulcer disease | K25, K26, K27, K28 | 0 |
| Mild liver disease | B18, K700, K701, K702, K703, K709, K713, K714, K715, K717, K73, K74, K760, K762, K763, K764, K768, K769, Z944 | 2 |
| Diabetes without chronic complication | E100, E101, E109, E110, E111, E119, E120, E121, E129, E130, E131, E139, E140, E141, E149 | 0 |
| Diabetes with chronic complication | E102, E103, E104, E105, E106, E107, E108, E112, E113, E114, E115, E116, E117, E118, E122, E123, E124, E125, E126, E127, E128, E132, E133, E134, E135, E136, E137, E138, E142, E143, E144, E145, E146, E147, E148 | 1 |
| Hemiplegia or paraplegia | G81, G82, G041, G114, G801, G802, G830, G831, G832, G833, G834, G839 | 2 |
| Renal disease | I120, I131, N032, N033, N034, N035, N036, N037, N052, N053, N054, N055, N056, N057, N18, N19, N250, Z490, Z491, Z492, Z940, Z992 | 1 |
| Any malignancy (including leukaemia and lymphoma) | C00, C01, C02, C03, C04, C05, C06, C07, C08, C09, C10, C11, C12, C13, C14, C15, C16, C17, C18, C19, C20, C21, C22, C23, C24, C25, C26, C30, C31, C32, C33, C34, C37, C38, C39, C40, C41, C43, C45, C46, C47, C48, C49, C50, C51, C52, C53, C54, C55, C56, C57, C58, C60, C61, C62, C63, C64, C65, C66, C67, C68, C69, C70, C71, C72, C73, C74, C75, C76, C81, C82, C83, C84, C85, C88, C90, C91, C92, C93, C94, C95, C96, C97 | 2 |
| Moderate or severe liver disease | I850, I859, I864, I982, K704, K711, K721, K729, K765, K766, K767 | 4 |
| Metastatic solid tumour | C77, C78, C79, C80 | 6 |
| AIDS / HIV | B20, B21, B22, B24 | 4 |

Charlson conditions and integer weights follow the update of Quan et al., Am J Epidemiol 2011;173:676–82. Where a Charlson condition shares its definition with an Elixhauser domain (congestive heart failure, peripheral vascular disease, chronic pulmonary disease, diabetes with and without complication, hemiplegia or paraplegia, metastatic cancer, AIDS/HIV), the identical code set is used. Mild and moderate-to-severe liver disease were scored additively rather than hierarchically, because Quan 2011 specifies no explicit hierarchy: a case coded for both therefore receives 2 plus 4 equals 6 liver points. All other conditions are independent flags.

The original Elixhauser comorbidity framework is described in Elixhauser et al., Med Care 1998;36:8–27. Aggregated outputs and the full computation code will be deposited on the Open Science Framework (DOI 10.17605/OSF.IO/P3QAW) upon publication.
