## Supplementary File S2: Pre-specified analysis code submitted to the Research Data Centre for "Do established comorbidity scores predict in-hospital mortality equally well in women and men? A nationwide validation in 166.7 million German inpatient cases, 2010–2024"

### Pre-specified analysis code submitted to the Research Data Centre

*Supplement to: Do established comorbidity scores predict in-hospital mortality equally well in women and men? A nationwide validation in 166.7 million German inpatient cases, 2010–2024*

Study registration: Open Science Framework, DOI 10.17605/OSF.IO/P3QAW

### What this file documents

The Research Data Centre of the Federal Statistical Office operates a controlled remote data processing model. The authors never access individual records: the complete analysis is written as a Stata do-file, submitted to the Research Data Centre, and executed there by Research Data Centre staff on the original data, and only aggregated output leaves the Centre. The submitted do-file therefore fixes the eligibility criteria, the comorbidity mappings and score definitions, the audit approach, the analysis sets, the age strata and the performance metrics before any result exists, in a form that cannot be revised afterwards. It is reproduced here so that the correspondence between the planned and the reported analysis can be checked directly. The do-file carries an internal creation date of 27 May 2026 and was submitted for execution before the aggregated outputs were returned.

Two submitted versions exist. They are identical in 1 036 of 1 046 lines. The differences are the switch of the execution environment from the local workstation to the Research Data Centre system, the file name for the 2024 reporting year once that file became available, the extension of the reporting-year loop from 2010–2023 to 2010–2024, a correction of the storage type of the case and event counters from int to long to prevent integer overflow at case counts in the millions, and header metadata. No eligibility criterion, comorbidity mapping, score definition, performance metric, analysis set, age stratum, pooling method or effect-size threshold differs between the two versions, and no aggregated output was released between them.

For data-protection reasons this excerpt begins with the study documentation and the analysis programs. The preceding environment block, which contains only Research Data Centre file paths, project identifiers and local directory settings, together with author contact details, is omitted; it contains no analytic content. Three comment lines carrying personal contact details and internal project labels have been removed, and one comment referring to an internal document has been replaced by a description of its content. No executable statement has been altered.

### Reproduced code

*Verbatim, 852 lines. Comments are partly in German as submitted; line numbers are those of the excerpt.*

```
* Purpose:
*   Audit of three established in-hospital comorbidity
*   scores (Charlson [Quan 2011 weights], Elixhauser sum, van Walraven score)
*   with respect to sex fairness, on the German DRG-Statistik.
*

* Cohort definition (per admission):
*     Step 1: Adults (alter >= 18, <= 110)
*     Step 2: Vollstationär (aufn_grd in {1, 2})
*     Step 3: No transfers IN  (aufn_anl not in {"V","A","K"})
*     Step 4: No transfers OUT (entl_grd not in {6, 8, 13, 29})
*     Step 5: Verweildauer >= 2 Belegungstage (tage >= 2)
*     Step 6: Sex known (sex in {"m","w"})
```

```
*     Step 7: Not pregnancy/childbirth (icd_hd not starting with "O")
*     Step 8: Not oncological therapy admission (icd_hd not starting with "Z51")
*     Step 9: Not in pseudo-departments (fab1 not in {0001-0006, xx60/xx61 Tagesklinik})
* Primary outcome: In-hospital mortality (entl_grd == 7).

* Sensitivity analyses (additional subsets of the main cohort):
*     "noSexCa"     - narrow: exclude only Mamma (C50) + Prostata (C61) as primary diagnosis
*     "noSexCancer" - medium: exclude all sex-specific oncology + benign tumors
*                     (C50-C58, C60-C63, D05-D07, D24-D29, D40)
*     "noSexSpec"   - broad: exclude all sex-specific diagnoses (noSexCancer plus
*                     N40-N51 male genital, N60-N64 mamma, N70-N98 female genital,
*                     E28.2 PCOS, E29 testicular dysfunction)
*     "emergency"   - restrict to emergency admissions (aufn_anl == "N")
*     "elective"    - restrict to elective admissions (aufn_anl in "E","Z")
*     "incl_preg"   - reverse: re-include pregnancy cases (Steps 1-6 + 8 + 9 only)

* Output: ONE workbook per year, output_phase1_YYYY.xlsx, with sheets:
*     T0_flowchart                Cohort exclusion tracking
*     T1_baseline                 Demographics + comorbidity prevalences by sex
*     T1b_baseline_notfall        Stratified by emergency/elective × sex
*     T1c_icdchapter              Frequency of ICD-10 main chapter × sex
*     T2_scoredist_<score>        Score value frequencies (n >= 100)
*     T3_mortality_<score>        Mortality by score quintile × sex
*     T4_T5b_<tag>                AUC + calibration metrics (HL, Spiegelhalter,
*                                 Brier, cal-in-large, slope) per analysis_set tag
*                                 (main, noSexCa, noSexCancer, noSexSpec, emergency,
*                                 elective, incl_preg)
*     T4b_aucdiff_<tag>           AUC-difference Z-test male vs female per tag
*     T5_calib_<score>_sex<sx>    Decile-aggregated predicted vs observed (main cohort)
*     T6_auc_byAge                AUC by age group × sex × score (main cohort)
*     T7_decisioncurve            Net Benefit at clinical thresholds (main cohort)


*******************************************************
*******************************************************



*******************************************************

program define audit_analysis
    syntax , year(integer) tag(string) outpath(string)

    tempname auc
    tempfile aucfile
    postfile `auc' int year byte sex_n str10 score ///
            long n_obs long n_events ///
            double auc se ci_lo ci_hi ///
            double cal_in_large cal_slope brier ///
            double hl_chi2 hl_df hl_p sp_z sp_p ///
            using `aucfile', replace

    foreach sc in cci elix vws {

        capture drop _p_pool _logitp _br_tmp _spnum _spden _dec_pool

        * 1) Gender-neutral pool-logit: dead ~ score on FULL current data
        capture quietly logit dead `sc'
        if _rc == 0 {

            * 2) Pool predicted probabilities
            quietly predict double _p_pool if e(sample), pr

            * 3) Sex-stratified metrics on the pooled predictions
            forvalues sx = 1/2 {
                quietly count if sex_n == `sx' & !missing(_p_pool)
                local n_sub = r(N)
                quietly count if sex_n == `sx' & !missing(_p_pool) & dead == 1
                local n_dead = r(N)

                if `n_sub' >= 100 {
```

```
* 3a) AUC + SE within sex subset
local auc_val = .
local se_val = .
capture quietly roctab dead _p_pool if sex_n == `sx'
if _rc == 0 {
    local auc_val = r(area)
    local se_val  = r(se)
}

* 3b) Calibration-in-the-Large
quietly summarize dead if sex_n == `sx' & !missing(_p_pool)
local obs_mean = r(mean)
quietly summarize _p_pool if sex_n == `sx'
local pred_mean = r(mean)
local cil = `obs_mean' - `pred_mean'

* 3c) Calibration-Slope: dead ~ logit(_p_pool) on sex subset
local slope = .
capture drop _logitp
quietly gen double _logitp = logit(_p_pool) if sex_n == `sx' & _p_pool > 0 & _p_pool < 1
capture quietly logit dead _logitp if sex_n == `sx'
if _rc == 0 local slope = _b[_logitp]
capture drop _logitp

* 3d) Hosmer-Lemeshow manuell: 10 Deciles auf Sex-Subset
*     mit Pool-Predictions. Robust: zählt nur valide Deciles
*     (n>=5 und 0<pi<1), passt df entsprechend an.
capture drop _dec_pool
quietly xtile _dec_pool = _p_pool if sex_n == `sx', n(10)
local hl_chi2 = 0
local hl_ndec = 0
forvalues d = 1/10 {
    quietly count if sex_n == `sx' & _dec_pool == `d'
    local n_d = r(N)
    if `n_d' >= 5 {
        quietly summarize dead if sex_n == `sx' & _dec_pool == `d', meanonly
        local O = r(sum)
        quietly summarize _p_pool if sex_n == `sx' & _dec_pool == `d', meanonly
        local pi = r(mean)
        if `pi' > 0 & `pi' < 1 {
            local E = `pi' * `n_d'
            local denom = `pi' * (1 - `pi') * `n_d'
            local hl_chi2 = `hl_chi2' + ((`O' - `E')^2) / `denom'
            local hl_ndec = `hl_ndec' + 1
        }
    }
}
capture drop _dec_pool
local hl_df = .
local hl_p = .
if `hl_ndec' >= 3 {
    local hl_df = `hl_ndec' - 2
    local hl_p  = 1 - chi2(`hl_df', `hl_chi2')
}
else {
    local hl_chi2 = .
}

* 3e) Spiegelhalter-Z
local sp_z = .
local sp_p = .
capture drop _spnum _spden
quietly gen double _spnum = (dead - _p_pool)*(1 - 2*_p_pool) if sex_n == `sx' & !missing(_p_pool)
quietly gen double _spden = ((1 - 2*_p_pool)^2) * _p_pool * (1 - _p_pool) if sex_n == `sx' & !missing(_p_pool)
quietly summarize _spnum, meanonly
local sp_num = r(sum)
quietly summarize _spden, meanonly
local sp_den_sq = r(sum)
```

```
                    if `sp_den_sq' > 0 {
                        local sp_z = `sp_num' / sqrt(`sp_den_sq')
                        local sp_p = 2*(1 - normal(abs(`sp_z')))
                    }
                    capture drop _spnum _spden

                    * 3f) Brier
                    capture drop _br_tmp
                    quietly gen double _br_tmp = (dead - _p_pool)^2 if sex_n == `sx'
                    quietly summarize _br_tmp if sex_n == `sx', meanonly
                    local brier_val = r(mean)
                    capture drop _br_tmp

                    * 3g) Post the row
                    post `auc' (`year') (`sx') ("`sc'") ///
                        (`n_sub') (`n_dead') ///
                        (`auc_val') (`se_val') ///
                        (`auc_val' - 1.96*`se_val') (`auc_val' + 1.96*`se_val') ///
                        (`cil') (`slope') (`brier_val') ///
                        (`hl_chi2') (`hl_df') (`hl_p') ///
                        (`sp_z') (`sp_p')
                }
            }
            capture drop _p_pool
        }
    }
    postclose `auc'

    * Export T4/T5b combined metrics
    preserve
        use `aucfile', clear
        gen str20 analysis_set = "`tag'"
        order analysis_set year sex_n score n_obs n_events ///
              auc se ci_lo ci_hi cal_in_large cal_slope brier ///
              hl_chi2 hl_df hl_p sp_z sp_p
        export excel using "`outpath'/output_phase1_`year'.xlsx", ///
               sheet("T4_T5b_`tag'") sheetreplace firstrow(variables)
    restore

    * T4b: AUC-Difference Z-Test (independent-sample DeLong equivalent)
    preserve
        use `aucfile', clear
        keep year sex_n score n_obs auc se
        reshape wide n_obs auc se, i(year score) j(sex_n)
        capture confirm variable auc1
        if _rc == 0 {
            capture confirm variable auc2
            if _rc == 0 {
                gen double auc_diff = auc1 - auc2
                gen double diff_se  = sqrt(se1^2 + se2^2)
                gen double z_diff   = auc_diff / diff_se
                gen double p_diff   = 2*(1 - normal(abs(z_diff)))
                rename n_obs1 n_male
                rename n_obs2 n_female
                rename auc1   auc_male
                rename auc2   auc_female
                rename se1    se_male
                rename se2    se_female
                gen str20 analysis_set = "`tag'"
                order analysis_set year score n_male n_female ///
                      auc_male se_male auc_female se_female ///
                      auc_diff diff_se z_diff p_diff
                export excel using "`outpath'/output_phase1_`year'.xlsx", ///
                       sheet("T4b_aucdiff_`tag'") sheetreplace firstrow(variables)
            }
        }
    restore
end
```

```
*    ------------------------------------------------------------------------------------------------------------
*    Subroutine 2: audit_analysis_byage
*    Age-stratified AUC on pool-logit predictions
*    ------------------------------------------------------------------------------------------------------------

program define audit_analysis_byage
    syntax , year(integer) outpath(string)

    tempname auca
    tempfile aucafile
    postfile `auca' int year byte agegrp byte sex_n str10 score ///
            long n_obs long n_events double auc se ///
            using `aucafile', replace

    foreach sc in cci elix vws {
        capture drop _p_age
        capture quietly logit dead `sc'
        if _rc == 0 {
            quietly predict double _p_age if e(sample), pr
            forvalues ag = 1/4 {
                forvalues sx = 1/2 {
                    quietly count if sex_n == `sx' & agegrp == `ag' & !missing(_p_age)
                    local n_sub = r(N)
                    quietly count if sex_n == `sx' & agegrp == `ag' & !missing(_p_age) & dead == 1
                    local n_dead = r(N)
                    if `n_sub' >= 100 {
                        capture quietly roctab dead _p_age if sex_n == `sx' & agegrp == `ag'
                        if _rc == 0 {
                            post `auca' (`year') (`ag') (`sx') ("`sc'") ///
                                         (`n_sub') (`n_dead') (r(area)) (r(se))
                        }
                    }
                }
            }
            capture drop _p_age
        }
    }
    postclose `auca'

    preserve
        use `aucafile', clear
        export excel using "`outpath'/output_phase1_`year'.xlsx", ///
               sheet("T6_auc_byAge") sheetreplace firstrow(variables)
    restore
end


*    ------------------------------------------------------------------------------------------------------------
*    Subroutine 3: audit_decision_curve  – Net Benefit at clinical thresholds
*    ------------------------------------------------------------------------------------------------------------

program define audit_decision_curve
    syntax , year(integer) outpath(string)

    tempname dc
    tempfile dcfile
    postfile `dc' int year byte sex_n str10 score ///
            double threshold double net_benefit long n_obs ///
            using `dcfile', replace

    foreach sc in cci elix vws {
        capture drop _p_dc
        capture quietly logit dead `sc'
        if _rc == 0 {
            quietly predict double _p_dc if e(sample), pr
            forvalues sx = 1/2 {
                quietly count if sex_n == `sx' & !missing(_p_dc)
```

```
                local n_total = r(N)
                if `n_total' >= 100 {
                    foreach t in 0.025 0.050 0.075 0.100 0.150 0.200 0.250 0.300 {
                        quietly count if sex_n == `sx' & _p_dc >= `t' & dead == 1
                        local tp = r(N)
                        quietly count if sex_n == `sx' & _p_dc >= `t' & dead == 0
                        local fp = r(N)
                        local pt = `t'
                        local nb = (`tp' / `n_total') - (`fp' / `n_total') * (`pt' / (1 - `pt'))
                        post `dc' (`year') (`sx') ("`sc'") (`pt') (`nb') (`n_total')
                    }
                }
            }
            capture drop _p_dc
        }
    }
    postclose `dc'

    preserve
        use `dcfile', clear
        export excel using "`outpath'/output_phase1_`year'.xlsx", ///
               sheet("T7_decisioncurve") sheetreplace firstrow(variables)
    restore
end


*    --------------------------------------------------------------------------------------------------------------------
*    Hauptloop pro Berichtsjahr
*    --------------------------------------------------------------------------------------------------------------------

local keep_vars icd_hd icd_nd1-icd_nd89 sex alter aufn_anl aufn_grd entl_grd tage beatm fab1

forvalues y = 2010/2024 {

    *    **********************************************************************
    *    Hauptanalyse 2010-2024 (15 Jahre, post-G-DRG-Konsolidierung).
    *    **********************************************************************

    * Datenladen je nach Arbeitsumgebung:
    *   FDZ=0 (lokal, Datenstrukturfile): $datenpfad + $dateiname_YYYY
    *   FDZ=1 (KDFV): $datenpfad_drg + $datei_drg_YYYY (FDZ-Standardvariablen ab März 2026)
    *   FDZ=2 (GWAP): analog FDZ=1
    if $FDZ == 0 {
        use `keep_vars' using "$datenpfad/${dateiname_`y'}", clear
    }
    if $FDZ == 1 {
        use `keep_vars' using "$datenpfad_drg/${datei_drg_`y'}", clear
    }
    if $FDZ == 2 {
        use `keep_vars' using "$datenpfad_drg/${datei_drg_`y'}", clear
    }
    compress
    gen year = `y'


    *    ------------------------------------------------------------------
    *    Sex (analog Projekt 15)
    *    ------------------------------------------------------------------
    gen byte sex_n = .
    replace sex_n = 1 if sex=="m"
    replace sex_n = 2 if sex=="w"
    label define sexlbl 1 "Male" 2 "Female", replace
    label values sex_n sexlbl


    *    ------------------------------------------------------------------
    *    Robust numeric age (analog Projekt 15)
    *    ------------------------------------------------------------------
    capture drop age_num
```

```
capture confirm numeric variable alter
if _rc {
    tempvar _ages
    gen strL `_ages' = trim(alter)
    replace `_ages' = subinstr(`_ages', ",", ".", .)
    destring `_ages', gen(age_num) force
    drop `_ages'
}
else gen double age_num = alter
replace age_num = . if age_num<0 | age_num>120


*    -----------------------------------------------------------------
*    Outcome (DSB: entl_grd numerisch; 7 = Tod)
*    -----------------------------------------------------------------
capture confirm string variable entl_grd
local entl_grd_string = (_rc == 0)
if `entl_grd_string' {
    gen byte dead = (entl_grd == "07")
}
else {
    gen byte dead = (entl_grd == 7)
}
label var dead "In-hospital Mortalität"


*    -----------------------------------------------------------------
*    Altersgruppen, Beatmung
*    -----------------------------------------------------------------
gen byte agegrp = .
replace agegrp = 1 if age_num >= 18 & age_num <= 44
replace agegrp = 2 if age_num >= 45 & age_num <= 64
replace agegrp = 3 if age_num >= 65 & age_num <= 79
replace agegrp = 4 if age_num >= 80
label define agelbl 1 "18-44" 2 "45-64" 3 "65-79" 4 ">=80", replace
label values agegrp agelbl

capture confirm numeric variable beatm
if !_rc gen byte vent = (beatm > 0 & !missing(beatm))
else    gen byte vent = .


*    -----------------------------------------------------------------
*    Notfall / Elektiv (DSB: aufn_anl string)
*    -----------------------------------------------------------------
gen byte notfall = 0
gen byte elektiv = 0
capture confirm string variable aufn_anl
if !_rc {
    replace notfall = (aufn_anl == "N")
    replace elektiv = inlist(aufn_anl, "E", "Z")
}
label var notfall "Notfall-Aufnahme"
label var elektiv "Elektive Aufnahme"


*    -----------------------------------------------------------------
*    Verweildauer-Kategorien
*    -----------------------------------------------------------------
gen byte los_cat = .
capture confirm numeric variable tage
if !_rc {
    replace los_cat = 1 if tage >= 2 & tage <= 4
    replace los_cat = 2 if tage >= 5 & tage <= 14
    replace los_cat = 3 if tage >= 15
}
label define loslbl 1 "2-4 Tage" 2 "5-14 Tage" 3 ">=15 Tage", replace
label values los_cat loslbl


*    -----------------------------------------------------------------
```

```
*    ICD-10 Hauptkapitel (A-Z) der Hauptdiagnose
*    ----------------------------------------------------------------
gen str1 icd_chapter = substr(icd_hd, 1, 1)


*    ----------------------------------------------------------------
*    Komorbiditäten (Elixhauser-Quan 2005) und Scores
*    Methodisch identisch zu Projekt 15 (regexm über all_diag)
*    ----------------------------------------------------------------
egen all_diag = concat(icd_hd icd_nd*), punct(" ")

* RAM-Optimierung: ICD-Nebendiagnosen nach Konkatenation droppen.
* Spart ~89 String-Variablen.
drop icd_nd*

gen HRST    = regexm(all_diag, "I47|I48|I49|I441|I442|I443|I456|I459|R000|R001|R008|T821|Z450|Z950")
gen KV      = regexm(all_diag, "I05|I06|I07|I08|I34|I35|I36|I37|I38|I39|I091|I098|A520|Q230|Q231|Q232|Q233|Z952|Z953|Z954")
gen PCD     = regexm(all_diag, "I26|I27|I280|I288|I289")
gen PVD     = regexm(all_diag, "I70|I71|I731|I738|I739|I771|I790|I792|K551|K558|K559|Z958|Z959")
gen HYPERT  = regexm(all_diag, "I10")
gen HYPERTC = regexm(all_diag, "I11|I12|I13|I15")
gen HI      = regexm(all_diag, "I43|I50|I099|I110|I130|I132|I255|I420|I425|I426|I427|I428|I429|P290")
gen PAR     = regexm(all_diag, "G81|G82|G041|G114|G801|G802|G830|G831|G832|G833|G834|G839")
gen OND     = regexm(all_diag, "G10|G11|G12|G13|G20|G21|G22|G32|G35|G36|G37|G40|G41|G254|G255|G312|G318|G319|G931|G934|R470|R56")
gen CPD     = regexm(all_diag, "J40|J41|J42|J43|J44|J45|J46|J47|J60|J61|J62|J63|J64|J65|J66|J67|I278|I279|J684|J701|J703")
gen DIA     = regexm(all_diag, "E100|E101|E109|E110|E111|E119|E120|E121|E129|E130|E131|E139|E140|E141|E149")
gen DIAC    = regexm(all_diag, "E102|E103|E104|E105|E106|E107|E108|E112|E113|E114|E115|E116|E117|E118|E122|E123|E124|E125|E126|E127|E128|E132|E133|E134|E135|E136|E137|E138|E142|E143|E144|E145|E146|E147|E148")
gen HYPO    = regexm(all_diag, "E00|E01|E02|E03|E890")
gen CKD     = regexm(all_diag, "N18|N19|I120|I131|N250|Z490|Z491|Z492|Z940|Z992")
gen LIVER   = regexm(all_diag, "B18|I85|K70|K72|K73|K74|I864|I982|K711|K713|K714|K715|K717|K760|K762|K763|K764|K765|K766|K767|K768|K769|Z944")
gen PUC     = regexm(all_diag, "K257|K259|K267|K269|K277|K279|K287|K289")
gen AIDS    = regexm(all_diag, "B20|B21|B22|B24")
gen LYMPH   = regexm(all_diag, "C81|C82|C83|C84|C85|C88|C96|C900|C902")
gen MET     = regexm(all_diag, "C77|C78|C79|C80")
gen SOLID   = regexm(all_diag, "C00|C01|C02|C03|C04|C05|C06|C07|C08|C09|C10|C11|C12|C13|C14|C15|C16|C17|C18|C19|C20|C21|C22|C23|C24|C25|C26|C30|C31|C32|C33|C34|C37|C38|C39|C40|C41|C43|C45|C46|C47|C48|C49|C50|C51|C52|C53|C54|C55|C56|C57|C58|C59|C60|C61|C62|C63|C64|C65|C66|C67|C68|C69|C70|C71|C72|C73|C74|C75|C76|C97")
gen RA      = regexm(all_diag, "L940|L941|L943|M05|M06|M08|M30|M32|M33|M34|M35|M45|M120|M123|M310|M311|M312|M313|M461|M468|M469")
gen COAG    = regexm(all_diag, "D65|D66|D67|D68|D691|D693|D694|D695|D696")
gen OBE     = regexm(all_diag, "E66")
gen WL      = regexm(all_diag, "E40|E41|E42|E43|E44|E45|E46|R634|R64")
gen FLUID   = regexm(all_diag, "E86|E87|E222")
gen BLDLOSS = regexm(all_diag, "D500")
gen ANEMDEF = regexm(all_diag, "D51|D52|D53|D508|D509")
gen ALCA    = regexm(all_diag, "E52|F10|T51|G621|I426|K292|K700|K703|K709|Z502|Z714|Z721")
gen DRUGA   = regexm(all_diag, "F11|F12|F13|F14|F15|F16|F18|F19|Z715|Z722")
gen PS      = regexm(all_diag, "F20|F22|F23|F24|F25|F28|F29|F302|F312|F315")
gen DEP     = regexm(all_diag, "F32|F33|F204|F313|F314|F315|F341|F412|F432")

* Van Walraven (published point weights)
gen vws = (-2*ANEMDEF) + (-2*BLDLOSS) + (7*HI) + (3*CPD) + (3*COAG) + (-3*DEP) + (-7*DRUGA) + ///
          (11*LIVER) + (9*LYMPH) + (5*FLUID) + (12*MET) + (6*OND) + (-4*OBE) + (7*PAR) + (4*PCD) + ///
          (2*PVD) + (5*CKD) + (4*SOLID) + (-1*KV) + (6*WL) + (5*HRST)

* Elixhauser-Sum
gen elix = HRST + KV + PCD + PVD + HYPERT + HYPERTC + HI + PAR + OND + CPD + DIA + DIAC + HYPO + CKD + LIVER + PUC + ///
           AIDS + LYMPH + MET + SOLID + RA + COAG + OBE + WL + FLUID + BLDLOSS + ANEMDEF + ALCA + DRUGA + PS + DEP

*    ----------------------------------------------------------------
*    Charlson Comorbidity Index nach Quan 2011 (eigenständige Definitionen,
```

```
*    nicht aus Elixhauser-Variablen geliehen, weil sich mehrere Phänotypen
*    unterscheiden: Rheumatic, PUD, Renal, Liver mild/severe)
*
*    Quan H, Li B, Couris CM, et al. Updating and Validating the Charlson
*    Comorbidity Index and Score for Risk Adjustment in Hospital Discharge
*    Abstracts Using Data From 6 Countries. Am J Epidemiol. 2011;173(6):676.
*    -----------------------------------------------------------------

gen cci_mi          = regexm(all_diag, "I21|I22|I252")
gen cci_chf         = HI                                                       /* identisch mit Elixhauser */
gen cci_pvd         = PVD                                                      /* identisch mit Elixhauser */
gen cci_cvd         = regexm(all_diag, "G45|G46|H340|I60|I61|I62|I63|I64|I65|I66|I67|I68|I69")
gen cci_dementia    = regexm(all_diag, "F00|F01|F02|F03|F051|G30|G311")
gen cci_copd        = CPD                                                      /* identisch mit Elixhauser CPD */
gen cci_rheum       = regexm(all_diag, "M05|M06|M315|M32|M33|M34|M351|M353|M360")
gen cci_pud         = regexm(all_diag, "K25|K26|K27|K28")                      /* alle Subcodes K25-K28 */
gen cci_liver_mild  = regexm(all_diag, "B18|K700|K701|K702|K703|K709|K713|K714|K715|K717|K73|K74|K760|K762|K763|K764|K768|K769|Z944")
gen cci_dm          = DIA                                                      /* identisch Elixhauser DM uncomplicated */
gen cci_dmcx        = DIAC                                                     /* identisch Elixhauser DM complicated */
gen cci_para        = PAR                                                      /* identisch Elixhauser */
gen cci_renal       = regexm(all_diag, "I120|I131|N032|N033|N034|N035|N036|N037|N052|N053|N054|N055|N056|N057|N18|N19|N250|Z490|Z491|Z492|Z940|Z992")
gen cci_cancer      = regexm(all_diag, "C00|C01|C02|C03|C04|C05|C06|C07|C08|C09|C10|C11|C12|C13|C14|C15|C16|C17|C18|C19|C20|C21|C22|C23|C24|C25|C26|C30|C31|C32|C33|C34|C37|C38|C39|C40|C41|C43|C45|C46|C47|C48|C49|C50|C51|C52|C53|C54|C55|C56|C57|C58|C60|C61|C62|C63|C64|C65|C66|C67|C68|C69|C70|C71|C72|C73|C74|C75|C76|C81|C82|C83|C84|C85|C88|C90|C91|C92|C93|C94|C95|C96|C97")
gen cci_liver_sev   = regexm(all_diag, "I850|I859|I864|I982|K704|K711|K721|K729|K765|K766|K767")
gen cci_mets        = MET                                                      /* identisch Elixhauser */
gen cci_aids        = AIDS                                                     /* identisch Elixhauser */

* Charlson-Score mit Quan 2011-Gewichten (6-Länder-Update):
*   MI 0, CHF 2, PVD 0, CVD 0, Dementia 2, COPD 1, Rheumatic 1, PUD 0,
*   Mild Liver 2, DM 0, DM cx 1, Hemiplegia 2, Renal 1, Any malignancy 2,
*   Mod/Sev Liver 4, Metastatic 6, AIDS 4.
* Hinweis: bei gleichzeitigem Mild- und Mod/Sev-Liver wird das schwerere
* gewichtet (Mod/Sev hat 4 Pkt vs. Mild 2 Pkt) - hier additiv gerechnet,
* weil Quan 2011 keine explizite Hierarchie spezifiziert. Mod/Sev Liver
* subsumiert konzeptionell die milde Variante (Liver-Disease = max-Schweregrad).
gen cci = (0*cci_mi) + (2*cci_chf) + (0*cci_pvd) + (0*cci_cvd) + (2*cci_dementia) + ///
          (1*cci_copd) + (1*cci_rheum) + (0*cci_pud) + ///
          (2*cci_liver_mild) + (0*cci_dm) + (1*cci_dmcx) + (2*cci_para) + ///
          (1*cci_renal) + (2*cci_cancer) + (4*cci_liver_sev) + (6*cci_mets) + (4*cci_aids)


*    -----------------------------------------------------------------
*    Sensitivitäts-Flags (Hauptdiagnose)
*    -----------------------------------------------------------------

* Schwangerschaft / Geburt / Wochenbett (Kapitel O als Hauptdiagnose)
gen byte preg_hd = regexm(icd_hd, "^O")

* Sex-spezifische Karzinome als Hauptdiagnose: Mamma (C50) und Prostata (C61)
gen byte sexca_hd = regexm(icd_hd, "^C50") | regexm(icd_hd, "^C61")

* Erweiterung: alle sex-spezifischen onkologischen Hauptdiagnosen
*   F: C50 Mamma, C51 Vulva, C52 Vagina, C53 Cervix, C54 Corpus,
*      C55 Uterus o.n.A., C56 Ovar, C57 Adnex, C58 Placenta,
*      D05 Mamma in situ, D06 Cervix in situ, D07.0-D07.3 weibl. Genital in situ,
*      D24 Mamma benigne, D25 Uterusmyom, D26-D28 weibl. Genital benigne
*   M: C60 Penis, C61 Prostata, C62 Hoden, C63 sonst. männl. Genital,
*      D07.4-D07.6 männl. Genital in situ, D29 männl. Genital benigne,
*      D40 männl. Genital unsicher
gen byte sexcancer_hd = ///
    regexm(icd_hd, "^C5[0-8]") | regexm(icd_hd, "^C6[0-3]") | ///
    regexm(icd_hd, "^D05") | regexm(icd_hd, "^D06") | ///
```

```
    regexm(icd_hd, "^D07[0-6]") | ///
    regexm(icd_hd, "^D2[4-9]") | ///
    regexm(icd_hd, "^D40")

* Maximaler Filter: alle sex-spezifischen Hauptdiagnosen (Onko + nicht-Onko)
*   Zusätzlich zu sexcancer_hd:
*     M: N40-N51 (männliche Genitalerkrankungen)
*     F: N60-N64 (Mamma), N70-N98 (weibliche Genital, Klimakterium, Infertilität)
*     E28.2 PCOS, E29 Hodenfunktionsstörung
gen byte sexspec_hd = sexcancer_hd | ///
    regexm(icd_hd, "^N4[0-9]") | regexm(icd_hd, "^N5[01]") | ///
    regexm(icd_hd, "^N6[0-4]") | ///
    regexm(icd_hd, "^N7[0-9]") | regexm(icd_hd, "^N8[0-9]") | ///
    regexm(icd_hd, "^N9[0-8]") | ///
    regexm(icd_hd, "^E282") | regexm(icd_hd, "^E29")

* Onkologische Therapie-Aufnahmen (Z51 Hauptdiagnose: Chemo, Strahlentherapie etc.)
gen byte z51_hd = regexm(icd_hd, "^Z51")

* Pseudo-Fachabteilungen (Tagesklinik / Stationsäquivalent etc.)
gen byte pseudo_fab = 0
capture confirm string variable fab1
if !_rc {
    replace pseudo_fab = 1 if inlist(fab1, "0001", "0002", "0003", "0004", "0005", "0006")
    replace pseudo_fab = 1 if regexm(fab1, "[0-9][0-9]6[01]$")
}
else {
    capture confirm numeric variable fab1
    if !_rc {
        replace pseudo_fab = 1 if inlist(fab1, 1, 2, 3, 4, 5, 6)
        replace pseudo_fab = 1 if mod(fab1, 100) == 60 | mod(fab1, 100) == 61
    }
}


*    -----------------------------------------------------------------
*    Kohortenfilter mit Flowchart-Tracking (T0)
*    -----------------------------------------------------------------
gen byte excl_step = 0
label define excl 0 "Eingeschlossen" ///
                  1 "Alter < 18 oder > 110" ///
                  2 "Nicht vollstationär (aufn_grd != 1, 2)" ///
                  3 "Verlegung in (aufn_anl in V, A, K)" ///
                  4 "Verlegung out (entl_grd in 6, 8, 13, 29)" ///
                  5 "Kurzzeitfall (< 2 Tage)" ///
                  6 "Geschlecht unbekannt" ///
                  7 "Schwangerschaft/Geburt (icd_hd in O)" ///
                  8 "Onkologische Therapie (icd_hd in Z51)" ///
                  9 "Pseudo-Fachabteilung", replace
label values excl_step excl

replace excl_step = 1 if age_num < 18 | age_num > 110 | missing(age_num)

capture confirm numeric variable aufn_grd
if !_rc replace excl_step = 2 if excl_step == 0 & !inlist(aufn_grd, 1, 2)

capture confirm string variable aufn_anl
if !_rc {
    replace excl_step = 3 if excl_step == 0 & inlist(aufn_anl, "V", "A", "K")
}

if `entl_grd_string' {
    replace excl_step = 4 if excl_step == 0 & inlist(entl_grd, "06", "08", "13", "29")
}
else {
    replace excl_step = 4 if excl_step == 0 & inlist(entl_grd, 6, 8, 13, 29)
}

capture confirm numeric variable tage
if !_rc replace excl_step = 5 if excl_step == 0 & tage < 2 & !missing(tage)
replace excl_step = 6 if excl_step == 0 & !inlist(sex_n, 1, 2)
```

```
    replace excl_step = 7 if excl_step == 0 & preg_hd == 1
    replace excl_step = 8 if excl_step == 0 & z51_hd == 1
    replace excl_step = 9 if excl_step == 0 & pseudo_fab == 1

    * Frische XLSX pro Jahr: alte ggf. löschen (sheetreplace würde sonst nur Sheets ersetzen, alte könnten von vorherigen Läufen mit weniger Sensitivitäten bleiben)
    capture rm "$outputpfad/output_phase1_`y'.xlsx"

    * Flowchart export
    preserve
        keep year excl_step
        contract year excl_step
        rename _freq n
        export excel using "$outputpfad/output_phase1_`y'.xlsx", ///
               sheet("T0_flowchart") sheetreplace firstrow(variables)
    restore


    *    ---------------------------------------------------------------
    *    Vor dem Drop: Snapshot speichern für incl_preg-Sensitivität
    *    ---------------------------------------------------------------
    tempfile snap_pre_main
    save `snap_pre_main', replace


    *    ---------------------------------------------------------------
    *    HAUPTKOHORTE: alle Filter anwenden
    *    ---------------------------------------------------------------
    keep if excl_step == 0
    drop excl_step

    gen byte one = 1


    *    ---------------------------------------------------------------
    *    T1: Baseline-Tabelle (Hauptkohorte) inkl. Notfall/Elektiv/LOS
    *    ---------------------------------------------------------------
    preserve
        collapse (sum) n_total = one ///
                 (mean) mort_rate = dead vent_rate = vent ///
                        mean_age = age_num ///
                        share_notfall = notfall share_elektiv = elektiv ///
                        share_HRST = HRST share_KV = KV share_PCD = PCD share_PVD = PVD ///
                        share_HYPERT = HYPERT share_HYPERTC = HYPERTC share_HI = HI ///
                        share_PAR = PAR share_OND = OND share_CPD = CPD ///
                        share_DIA = DIA share_DIAC = DIAC share_HYPO = HYPO ///
                        share_CKD = CKD share_LIVER = LIVER share_PUC = PUC ///
                        share_AIDS = AIDS share_LYMPH = LYMPH share_MET = MET share_SOLID = SOLID ///
                        share_RA = RA share_COAG = COAG share_OBE = OBE share_WL = WL ///
                        share_FLUID = FLUID share_BLDLOSS = BLDLOSS share_ANEMDEF = ANEMDEF ///
                        share_ALCA = ALCA share_DRUGA = DRUGA share_PS = PS share_DEP = DEP ///
                        share_sexca = sexca_hd ///
                 (mean) mean_cci = cci mean_elix = elix mean_vws = vws ///
                 (sd)   sd_cci = cci sd_elix = elix sd_vws = vws ///
                 (mean) mean_los = tage ///
                 (p50)  median_los = tage, ///
                 by(year sex_n)
        export excel using "$outputpfad/output_phase1_`y'.xlsx", ///
               sheet("T1_baseline") sheetreplace firstrow(variables)
    restore

    * T1b: Baseline-Tabelle nach Notfall/Elektiv × Sex (zusätzliche Stratifizierung)
    preserve
        gen byte category = .
        replace category = 1 if notfall == 1
        replace category = 2 if elektiv == 1
        replace category = 3 if notfall == 0 & elektiv == 0
        label define catlbl 1 "Notfall" 2 "Elektiv" 3 "Sonstiges", replace
        label values category catlbl
        collapse (sum) n_total = one ///
                 (mean) mort_rate = dead mean_age = age_num mean_los = tage ///
```

```
                    mean_cci = cci mean_elix = elix mean_vws = vws, ///
             by(year sex_n category)
    keep if n_total >= 100
    export excel using "$outputpfad/output_phase1_`y'.xlsx", ///
           sheet("T1b_baseline_notfall") sheetreplace firstrow(variables)
restore

* T1c: Verteilung Hauptdiagnose-Kapitel × Sex
preserve
    contract year sex_n icd_chapter
    rename _freq n
    keep if n >= 100
    export excel using "$outputpfad/output_phase1_`y'.xlsx", ///
           sheet("T1c_icdchapter") sheetreplace firstrow(variables)
restore


*    ----------------------------------------------------------------
*    T2: Score-Verteilung
*    ----------------------------------------------------------------
foreach sc in cci elix vws {
    preserve
        keep year sex_n `sc'
        rename `sc' score_value
        gen str10 score_type = "`sc'"
        contract year sex_n score_type score_value
        rename _freq n
        keep if n >= 100
        export excel using "$outputpfad/output_phase1_`y'.xlsx", ///
               sheet("T2_scoredist_`sc'") sheetreplace firstrow(variables)
    restore
}


*    ----------------------------------------------------------------
*    T3: Mortalität nach Score-Quintil
*    ----------------------------------------------------------------
foreach sc in cci elix vws {
    preserve
        xtile q_`sc' = `sc', n(5)
        collapse (sum) n = one (mean) mort_rate = dead, by(year sex_n q_`sc')
        rename q_`sc' quintile
        gen str10 score = "`sc'"
        keep if n >= 100
        order year sex_n score quintile n mort_rate
        export excel using "$outputpfad/output_phase1_`y'.xlsx", ///
               sheet("T3_mortality_`sc'") sheetreplace firstrow(variables)
    restore
}


*    ----------------------------------------------------------------
*    HAUPTANALYSE: T4_T5b + T4b für die volle Hauptkohorte (tag="main")
*    ----------------------------------------------------------------
audit_analysis, year(`y') tag("main") outpath("$outputpfad")


*    ----------------------------------------------------------------
*    T5: Decile-Calibration auf Pool-Predictions (Hauptkohorte)
*    ----------------------------------------------------------------
foreach sc in cci elix vws {
    capture drop _p_t5
    capture quietly logit dead `sc'
    if _rc == 0 {
        quietly predict double _p_t5 if e(sample), pr
        forvalues sx = 1/2 {
            capture drop _dec_t5
            capture quietly xtile _dec_t5 = _p_t5 if sex_n == `sx', n(10)
            if _rc == 0 {
                preserve
                    keep if sex_n == `sx' & !missing(_dec_t5)
```

```
                    collapse (sum) n = one (mean) mean_pred = _p_t5 obs_rate = dead, ///
                             by(year sex_n _dec_t5)
                    rename _dec_t5 decile
                    gen str10 score = "`sc'"
                    keep if n >= 100
                    order year sex_n score decile n mean_pred obs_rate
                    export excel using "$outputpfad/output_phase1_`y'.xlsx", ///
                           sheet("T5_calib_`sc'_sex`sx'") sheetreplace firstrow(variables)
                restore
            }
            capture drop _dec_t5
        }
        capture drop _p_t5
    }
}


*    ---------------------------------------------------------------
*    T6: Altersstratifizierte AUC
*    ---------------------------------------------------------------
audit_analysis_byage, year(`y') outpath("$outputpfad")


*    ---------------------------------------------------------------
*    T7: Decision Curve Analysis
*    ---------------------------------------------------------------
audit_decision_curve, year(`y') outpath("$outputpfad")


*    ---------------------------------------------------------------
*    SENSITIVITÄT 1a: noSexCa  (eng - nur Mamma C50 + Prostata C61 als HD raus)
*    ---------------------------------------------------------------
preserve
    drop if sexca_hd == 1
    audit_analysis, year(`y') tag("noSexCa") outpath("$outputpfad")
restore

*    ---------------------------------------------------------------
*    SENSITIVITÄT 1b: noSexCancer  (mittel - alle sex-spez. Onkologie als HD raus)
*    ---------------------------------------------------------------
preserve
    drop if sexcancer_hd == 1
    audit_analysis, year(`y') tag("noSexCancer") outpath("$outputpfad")
restore

*    ---------------------------------------------------------------
*    SENSITIVITÄT 1c: noSexSpec  (breit - alle sex-spez. Erkrankungen als HD raus)
*    ---------------------------------------------------------------
preserve
    drop if sexspec_hd == 1
    audit_analysis, year(`y') tag("noSexSpec") outpath("$outputpfad")
restore


*    ---------------------------------------------------------------
*    SENSITIVITÄT 2: emergency-only
*    ---------------------------------------------------------------
preserve
    keep if notfall == 1
    audit_analysis, year(`y') tag("emergency") outpath("$outputpfad")
restore


*    ---------------------------------------------------------------
*    SENSITIVITÄT 3: elective-only
*    ---------------------------------------------------------------
preserve
    keep if elektiv == 1
    audit_analysis, year(`y') tag("elective") outpath("$outputpfad")
restore
```

```
    *    ----------------------------------------------------------------
    *    SENSITIVITÄT 4: incl_preg (Schwangerschaft drin, sonst alle Filter)
    *    Snapshot von vor Hauptfilter holen, Schwangerschaft EINSCHLIESSEN
    *    ----------------------------------------------------------------
    use `snap_pre_main', clear
    * Alle Hauptkohorten-Filter, AUSSER Step 7 (Schwangerschaft):
    keep if inlist(excl_step, 0, 7)
    drop excl_step
    gen byte one = 1
    audit_analysis, year(`y') tag("incl_preg") outpath("$outputpfad")

}

* Timer beenden und Laufzeit ausgeben (FDZ-Standard)
timer off 1
timer list 1

log close
```
