## Supplementary File S3: Extended methodological considerations for "Do established comorbidity scores predict in-hospital mortality equally well in women and men? A nationwide validation in 166.7 million German inpatient cases, 2010–2024"

### Extended methodological considerations

*Supplement to: Do established comorbidity scores predict in-hospital mortality equally well in women and men? A nationwide validation in 166.7 million German inpatient cases, 2010–2024*

Study registration: Open Science Framework, DOI 10.17605/OSF.IO/P3QAW

Reference numbers correspond to the reference list of the main manuscript.

### Purpose

The Discussion of the main manuscript summarises the limitations that bear directly on the reported findings. This supplement sets out the full set of methodological considerations that bound the interpretation and the generalisability of the analysis, in the order in which they are referred to in the Discussion. Each item states the issue, the available evidence, the consequence for the present findings and, where applicable, the planned mitigation.

#### 1. Score construction and the original outcome of each score

Of the three scores examined, only the van Walraven Score was originally developed with in-hospital mortality as its native outcome [12]. The Charlson Comorbidity Index was developed in 559 medical inpatients with 1-year mortality as the outcome and validated over 10 years in 685 women treated for breast cancer [8]; its use for in-hospital mortality rests on the later re-weighting by Quan and colleagues [9]. The Elixhauser Comorbidity Sum is an unweighted count and is therefore not weighted by how strongly each condition predicts death. That the direction of the slope deviation is opposite between Charlson and Elixhauser-Sum, although both were computed from identical records with an identical fitting procedure, locates the difference in how each score selects and weights comorbidity domains rather than in the underlying data. The score-specific direction of the sex-related difference we report, namely Charlson and van Walraven favouring women and Elixhauser-Sum favouring men, may therefore reflect not only differences in the empirical sex-by-score-mortality relationship but also the developmental heterogeneity of the score constructions themselves. A formal test of this hypothesis, namely re-fitting all three scores with sex-stratified weights on the same German cohort and re-running the sex-stratified evaluation, is planned follow-up work that will test whether the score-specific direction of the sex difference persists after sex-stratified re-fitting. Both directions of result (persistence and attenuation) are methodologically informative.

#### 2. Present-on-admission versus hospital-acquired comorbidity coding

Unlike US Medicare since 2008, German DRG coding does not systematically flag present-on-admission (POA) status for secondary diagnoses. This prevents reliable separation of pre-existing comorbidities from conditions arising during the hospital stay (including hospital-acquired complications and incidentally documented conditions). Pine and colleagues reported in JAMA 2007 [33] that adding POA indicators to risk-adjustment models meaningfully changed hospital risk-adjusted mortality rankings; Glance and colleagues confirmed this in 2007 [34]. In our German DRG cohort, some "comorbidities" counted toward the three scores are likely in-hospital acquired complications. Fluid/electrolyte disorders, coagulopathy, and certain acute neurological events are particularly suspect. This introduces partial reverse causation: A complication causes both score-elevation and death, with the score-elevation appearing post hoc to predict the death it was caused by. All three scores in this study share this POA-ambiguity equally (it is a data-source feature, not a score-construction feature), so the between-score contrast in our headline findings is robust to it. The absolute calibration finding for any one score, however, is partially confounded by the POA-vs-HAC mixture. Whether the sex-related pattern is itself affected by sex-differences in complication-incidence is unclear without POA-stratified data; men have higher prevalence of some comorbidities suspect of post-admission coding inflation (mechanical ventilation 2.54 % vs 1.67 %, coagulopathy 4.02 % vs 3.02 %), whereas women have higher prevalence of others (fluid and electrolyte disorders 19.84 % vs 16.88 %). The net direction of any sex-related hospital-acquired-coding bias therefore cannot be signed without present-on-admission data and remains indeterminate.

#### 3. Documentation versus clinical effects

Three of the van Walraven score components, namely depression (penalty −3), drug abuse (penalty −7), and obesity (penalty −4), are documentation-sensitive in a sex-related way. In our cohort, depression was coded in 6.92 % of women's admissions versus 3.47 % of men's; obesity in 7.81 % versus 6.95 %. Whether these differences reflect a true clinical-prevalence difference or an asymmetric coding habit cannot be disentangled in administrative data alone. To the extent that documentation is asymmetric, the van Walraven score penalises women for a structural artefact of clinical coding habit rather than for an underlying mortality-relevant clinical condition. Our findings should be interpreted as an upper bound of a true-clinical sex-related difference effect and a lower bound of a structural-score-construction effect; sex-aware recalibration of the three scores must address both layers if it is to be robust to documentation drift.

#### 4. Single-admission comorbidity look-back

Single-admission look-back: Comorbidities are identified from the current admission only; Westerberg and colleagues showed that longitudinal look-back materially improves Charlson discrimination [17], and Schneeweiss reported analogous improvements in Medicare data [29]

#### 5. Hospital-level clustering

Hospital-level clustering: Our standard errors do not adjust for intra-hospital correlation (ICC ≈ 0.05–0.15 typically [30]), because we did not adjust for this clustering, our confidence intervals are likely somewhat too narrow; hospital identifiers are not contained in the aggregated FDZ outputs, so a cluster-robust re-estimation would require a separate Research Data Centre application and remains a limitation

#### 6. Linearity of the score-mortality relationship

Linearity of the score-mortality relationship: Some component of the slope deviation may reflect a linearity-violation × sex interaction, which a continuous-data restricted-cubic-spline analysis would directly test (planned as follow-up work)

#### 7. Cohort selection relative to German inpatient care

Cohort selection bias: The DRG-billed acute somatic cohort excludes psychiatric admissions (PEPP-billed; female-predominant) and rehabilitation admissions, biasing our female subpopulation toward the somatically-ill

### 8. End-of-life coding effect

End-of-life coding effect: Patients dying in hospital typically receive more diagnostic coding than equally-comorbid survivors, introducing reverse-causation; the sex-direction is unclear but the effect is shared between sexes

### 9. In-hospital versus 30-day mortality

In-hospital versus 30-day mortality: Drye and colleagues reported that the choice between in-hospital and 30-day outcomes shifts 10–15 % of hospital quintile classifications [35]; our findings are conditional on the in-hospital admission window. Three of these features (i, iii) potentially attenuate the score-component contribution. Three others (ii, v, vi) primarily affect uncertainty quantification. Taken together they constitute a substantial agenda for follow-up work. None of them changes the substantive calibration finding reported here.

### 10. Multiple testing

The evaluation covers three scores, four age strata, seven analysis sets and 15 reporting years on two performance axes. No formal multiple-testing correction is applied. Instead, effect-size thresholds were pre-specified before any result existed (between-sex calibration-slope difference of at least 0.05, absolute AUC difference of at least 0.01), and all sub-analyses are reported rather than selected. This follows the rationale that effect-size pre-specification with complete reporting is the appropriate discipline in large-scale descriptive epidemiology [25]. At the realised sample sizes, any Bonferroni-Holm or false-discovery-rate correction applied to the reported p-values would leave the set of effects exceeding the pre-specified thresholds unchanged.

### Summary of expected direction

Two considerations, the single-admission look-back and the end-of-life coding effect, would if anything attenuate rather than amplify the sex-related difference reported here, so the difference observed with a longitudinal look-back would plausibly be somewhat larger. Three considerations, hospital-level clustering, the linearity assumption and the in-hospital outcome window, affect the precision or the transportability of the estimates rather than the direction of the finding. The remaining considerations, score construction, present-on-admission coding, documentation practice and cohort selection, have directions that cannot be signed with the available data. None of them changes the direction of the calibration finding reported in the main manuscript.
