## Supplementary material for "Do established comorbidity scores predict in-hospital mortality equally well in women and men? A nationwide validation in 166.7 million German inpatient cases, 2010–2024": TRIPOD+AI Checklist

### TRIPOD+AI Checklist for the reporting of prediction model studies

Filled for: Sex-asymmetric calibration of three established comorbidity scores applied to in-hospital mortality (German DRG statistics 2010–2024)

*Official TRIPOD+AI checklist (www.tripod-statement.org, updated 23 April 2024). D = development-only item, E = evaluation-only, D;E = both. This study is an external evaluation/validation (TRIPOD type 4) of three previously published comorbidity scores; development-only (D) items are marked Not applicable. Section references point to manuscript HERA_Paper1_v1.8; final page/line numbers to be inserted from the typeset proof. Submit as a separate reporting-checklist document.*

| Topic | Item | D/E | Checklist item | Reported in (manuscript section) |
|---|---|---|---|---|
| **§ Title** | | | | |
| Title | 1 | D;E | Identify the study as developing or evaluating the performance of a multivariable prediction model, the target population, and the outcome to be predicted | Title |
| **§ Abstract** | | | | |
| Abstract | 2 | D;E | See TRIPOD+AI for Abstracts checklist | Abstract (see TRIPOD+AI for Abstracts below) |
| **§ Introduction** | | | | |
| Background | 3a | D;E | Explain the healthcare context (including whether diagnostic or prognostic) and rationale for developing or evaluating the prediction model, including references to existing models | Introduction |
| | 3b | D;E | Describe the target population and the intended purpose of the prediction model in the context of the care pathway, including its intended users | Introduction (risk adjustment and hospital quality reporting) |
| | 3c | D;E | Describe any known health inequalities between sociodemographic groups | Introduction (sex disparities; sex-asymmetric calibration of prior cohorts) |
| Objectives | 4 | D;E | Specify the study objectives, including whether the study describes the development or validation of a prediction model (or both) | Introduction (final paragraph); Abstract — Objectives (external validation, TRIPOD type 4) |
| **§ Methods** | | | | |
| Data | 5a | D;E | Describe the sources of data separately for the development and evaluation datasets, the rationale for using these data, and representativeness | Methods — Data source and cohort construction (German DRG statistics; complete national enumeration) |
| | 5b | D;E | Specify the dates of the collected participant data, including start and end of participant accrual; and, if applicable, end of follow-up | Methods — Data source (reporting years 2010 to 2024) |
| Participants | 6a | D;E | Specify key elements of the study setting including the number and location of centres | Methods — Data source (all German acute somatic DRG-billing hospitals, nationwide) |
| | 6b | D;E | Describe the eligibility criteria for study participants | Methods — Data source (nine-step eligibility filter); Figure 1 |
| | 6c | D;E | Give details of any treatments received, and how they were handled during model development or evaluation, if relevant | Not applicable (risk-adjustment performance audit; treatments not modelled) |
| Data preparation | 7 | D;E | Describe any data pre-processing and quality checking, including whether this was similar across relevant sociodemographic groups | Methods — Data source (per-step exclusion accounting); Statistical analysis; Verification, software |
| Outcome | 8a | D;E | Clearly define the outcome that is being predicted and the time horizon, including how and when assessed, the rationale, and whether assessment is consistent across sociodemographic groups | Methods — Outcome and predictors (in-hospital mortality, administrative discharge code, identical across sexes) |
| | 8b | D;E | If outcome assessment requires subjective interpretation, describe the qualifications and demographic characteristics of the outcome assessors | Not applicable (objective administrative discharge status; no subjective assessment) |
| | 8c | D;E | Report any actions to blind assessment of the outcome to be predicted | Not applicable (routinely collected administrative outcome) |
| Predictors | 9a | D | Describe the choice of initial predictors and any pre-selection of predictors before model building | Not applicable — external evaluation/validation of previously published scores; no model development in this study. |
| | 9b | D;E | Clearly define all predictors, including how and when they were measured | Methods — Outcome and predictors; Supplementary File S1 (ICD-10-GM code definitions for all score components) |
| | 9c | D;E | If predictor measurement requires subjective interpretation, describe the qualifications and demographic characteristics of the predictor assessors | Not applicable (predictors derived from coded ICD-10-GM diagnoses; Supplementary File S1) |
| Sample size | 10 | D;E | Explain how the study size was arrived at, and justify that it was sufficient | Methods — Data source (complete enumeration; n = 166 693 491); |

| | | | | |
|---|---|---|---|---|
| | | | | Abstract — Participants |
| Missing data | 11 | D;E | Describe how missing data were handled; provide reasons for omitting any data | Methods — Data source (sex-documented inclusion step); Statistical analysis |
| Analytical methods | 12a | D | Describe how the data were used (development and evaluation of performance), including any partitioning | Not applicable — external evaluation/validation of previously published scores; no model development in this study. |
| | 12b | D | Describe how predictors were handled in the analyses (functional form, rescaling, transformation, standardisation) | Not applicable — external evaluation/validation of previously published scores; no model development in this study. Score values used as published; see Supplementary File S1. |
| | 12c | D | Specify the type of model, rationale, all model building steps, and method for internal validation | Not applicable — external evaluation/validation of previously published scores; no model development in this study. |
| | 12d | D;E | Describe if and how heterogeneity in model parameter values and performance was handled and quantified across clusters | Methods — Statistical analysis (per-year DerSimonian-Laird random-effects pooling; $I^2$ heterogeneity across 15 reporting years) |
| | 12e | D;E | Specify all measures and plots used to evaluate model performance (eg, discrimination, calibration, clinical utility) | Methods — Statistical analysis and fairness assessment (CITL, calibration slope, decile calibration, Brier, AUC, observed mortality per quintile) |
| | 12f | E | Describe any model updating (eg, recalibration) arising from the model evaluation | No updating performed; sex-aware re-calibration discussed as planned follow-up (Discussion — Implications and research priorities) |
| | 12g | D;E | For model evaluation, describe how the model predictions were calculated (eg, formula, code) | Methods — Outcome and predictors; Supplementary File S1 (full score formulas and weights); Statistical analysis (pooled-logit predicted probabilities) |
| Class imbalance | 13 | D;E | If class imbalance methods were used, state why and how, and any recalibration | Not applicable (no class-imbalance resampling; full-population logistic models) |
| Fairness | 14 | D;E | Describe any approaches that were used to address model fairness and their rationale | Methods — Statistical analysis and fairness assessment (sex-stratified audit; pre-specified clinical-meaningfulness thresholds); SAGER 2016 adherence |
| Model output | 15 | D | Specify the output of the prediction model and details of any classification thresholds | Not applicable — external evaluation/validation of previously published scores; no model development in this study. Outcome is predicted in-hospital mortality probability. |
| Training vs evaluation | 16 | D;E | Identify any differences between the development and evaluation data in setting, eligibility, outcome, and predictors | External validation of three previously published scores (Charlson/Quan 2011, Elixhauser-Sum/Quan 2005, van Walraven 2009) on German DRG data; see Methods — Data source and Outcome and predictors |
| Ethical approval | 17 | D;E | Name the institutional research board or ethics committee that approved the study and describe consent or its waiver | Methods — Verification, software, reporting standards, ethics, PPI (§27 Federal Statistics Act; ethics approval not required for anonymised administrative data); Patient consent for publication |
| **§ Open science** | | | | |
| Funding | 18a | D;E | Give the source of funding and the role of the funders for the present study | Funding (None.) |
| Conflicts of interest | 18b | D;E | Declare any conflicts of interest and financial disclosures for all authors | Competing interests |
| Protocol | 18c | D;E | Indicate where the study protocol can be accessed or state that a protocol was not prepared | Methods — Study design and registration (OSF registration; retrospective — see Methods) |
| Registration | 18d | D;E | Provide registration information for the study, including register name and registration number | Abstract — Study registration; Methods — Study design and registration (OSF DOI 10.17605/OSF.IO/P3QAW) |
| Data sharing | 18e | D;E | Provide details of the availability of the study data | Data availability statement |
| Code sharing | 18f | D;E | Provide details of the availability of the analytical code | Data availability statement; Supplementary File S1 (score code definitions) |
| **§ Patient and public involvement** | | | | |
| PPI | 19 | D;E | Provide details of any patient and public involvement during design, conduct, reporting, | Methods — Verification, software, reporting standards, ethics, PPI |

| | | | | |
|---|---|---|---|---|
| | | | interpretation, or dissemination, or state no involvement | |
| **§ Results** | | | | |
| Participants | 20a | D;E | Describe the flow of participants through the study, including numbers with and without the outcome; a diagram may be helpful | Results — Cohort characteristics; Figure 1 (cohort flow diagram) |
| | 20b | D;E | Report the characteristics overall and, where applicable, for each data source or setting, including key predictors (including demographics), sample size, number of outcome events; report any differences across key demographic groups | Results — Cohort characteristics; Table 1 (by sex) |
| | 20c | E | For model evaluation, show a comparison with the development data of the distribution of important predictors | Not applicable (development-data distributions of the original score derivations not available; external validation only) |
| Model development | 21 | D;E | Specify the number of participants and outcome events in each analysis | Results — Cohort characteristics; Tables 1–8 (n and events per analysis/stratum) |
| Model specification | 22 | D | Provide details of the full prediction model to allow predictions in new individuals | Evaluated scores fully specified in Supplementary File S1 (components, ICD-10-GM codes, weights); no new model developed |
| Model performance | 23a | D;E | Report model performance estimates with confidence intervals, including for any key subgroups; consider plots | Results — Calibration by sex; Discrimination by sex; Tables 2–5, 7, 8; Figures 2–7 |
| | 23b | D;E | If examined, report results of any heterogeneity in model performance across clusters | Results — Discrimination by sex ($I^2$ across 15 reporting years); Figures 3, 5, 6 |
| Model updating | 24 | E | Report the results from any model updating, including the updated model and subsequent performance | Not applicable (no model updating performed in this study) |
| **§ Discussion** | | | | |
| Interpretation | 25 | D;E | Give an overall interpretation of the main results, including issues of fairness in the context of the objectives and previous studies | Discussion (Interpretation and subsequent subsections) |
| Limitations | 26 | D;E | Discuss any limitations of the study and their effects on biases, statistical uncertainty, and generalisability | Discussion — Strengths and limitations; Other methodological considerations |
| Usability | 27a | D | Describe how poor quality or unavailable input data should be assessed and handled when implementing the prediction model | Not applicable — external evaluation/validation of previously published scores; no model development in this study. |
| | 27b | D | Specify whether users will be required to interact in the handling of the input data or use of the model, and what expertise is required | Not applicable — external evaluation/validation of previously published scores; no model development in this study. |
| | 27c | D;E | Discuss any next steps for future research, with a specific view to applicability and generalisability of the model | Discussion — Implications and research priorities |

#### TRIPOD+AI for Abstracts

| Item | Checklist item | Reported in (manuscript section) |
|---|---|---|
| 1 | Identify the study as developing or evaluating the performance of a multivariable prediction model, the target population, and the outcome to be predicted | Title; Abstract — Objectives/Design |
| 2 | Provide a brief explanation of the healthcare context and rationale | Abstract — Objectives |
| 3 | Specify the study objectives, including whether the study describes model development, evaluation, or both | Abstract — Objectives/Design (evaluation; TRIPOD type 4) |
| 4 | Describe the sources of data | Abstract — Setting |
| 5 | Describe the eligibility criteria and setting where the data were collected | Abstract — Setting; Participants |
| 6 | Specify the outcome to be predicted, including time horizon for prognostic models | Abstract — Main outcome measures (in-hospital mortality) |
| 7 | Specify the type of model, summary of model-building steps, and method for internal validation | Not applicable (external validation of published scores) |
| 8 | Specify the measures used to assess model performance | Abstract — Main outcome measures (calibration; discrimination) |
| 9 | Report the number of participants and outcome events | Abstract — Participants/Results |

| | | |
|---|---|---|
| 10 | Summarise the predictors in the final model | Abstract — Objectives (three established scores; full definitions in Supplementary File S1) |
| 11 | Report model performance estimates (with confidence intervals) | Abstract — Results (slope, risk ratios, ΔAUC with CIs) |
| 12 | Give an overall interpretation of the main results | Abstract — Conclusions |
| 13 | Give the registration number and name of the registry or repository | Abstract — Study registration (OSF DOI 10.17605/OSF.IO/P3QAW) |
